# Optimizing COVID-19 surveillance in long-term care facilities: a modelling study

**DOI:** 10.1101/2020.04.19.20071639

**Authors:** David RM Smith, Audrey Duval, Koen B Pouwels, Didier Guillemot, Jérôme Fernandes, Bich-Tram Huynh, Laura Temime, Lulla Opatowski on behalf of the AP-HP/Universities/Inserm COVID-19 research collaboration

**Author notes:** contributed equally (David RM Smith, Audrey Duval). contributed equally (Laura Temime, Lulla Opatowski). Corresponding author: David R. M. Smith, 25 rue du Dr Roux Paris 75015 France.

## Abstract

**Background:** Long-term care facilities (LTCFs) are vulnerable to COVID-19 outbreaks. Timely epidemiological surveillance is essential for outbreak response, but is complicated by a high proportion of silent (non-symptomatic) infections and limited testing resources.

**Methods:** We used a stochastic, individual-based model to simulate SARS-CoV-2 transmission along detailed inter-individual contact networks describing patient-staff interactions in real LTCF settings. We distributed nasopharyngeal swabs and RT-PCR tests using clinical and demographic indications, and evaluated the efficacy and resource-efficiency of a range of surveillance strategies, including group testing (sample pooling) and testing cascades, which couple (i) testing for multiple indications (symptoms, admission) with (ii) random daily testing.

**Results:** In the baseline scenario, randomly introducing SARS-CoV-2 into a 170-bed LTCF led to large outbreaks, with a cumulative 86 (6-224) infections after three weeks of unmitigated transmission. Efficacy of symptom-based screening was limited by (i) lags between infection and symptom onset, and (ii) silent transmission from asymptomatic and pre-symptomatic infections. Testing upon admission detected up to 66% of patients silently infected upon LTCF entry, but missed potential introductions from staff. Random daily testing was more effective when targeting patients than staff, but was overall an inefficient use of limited resources. At high testing capacity (>1 test/10 beds/day), cascades were most effective, with a 22-52% probability of detecting outbreaks prior to any nosocomial transmission, and 38-63% prior to first onset of COVID-19 symptoms. Conversely, at low capacity (<1 test/85 beds/day), pooling randomly selected patients in a daily group test was most effective (9-15% probability of detecting outbreaks prior to transmission; 30-44% prior to symptoms). The most efficient strategy compared to the reference was to pool individuals with any COVID-like symptoms, requiring only 5-7 additional tests and 17-24 additional swabs to detect outbreaks 5-6 days earlier, prior to an additional 14-18 infections.

**Conclusions:** Group testing is an effective and efficient COVID-19 surveillance strategy for resource-limited LTCFs. Cascades are even more effective given ample testing resources. Increasing testing capacity and updating surveillance protocols accordingly could facilitate earlier detection of emerging outbreaks, informing a need for urgent intervention in settings with ongoing nosocomial transmission.

## BACKGROUND

From nursing homes to rehabilitation hospitals, long-term care facilities (LTCFs) are hotspots for COVID-19 outbreaks worldwide.(1) LTCF patients (or residents) require continuing care, live in close proximity to one another, and are typically elderly and multimorbid, placing them at elevated risk both of acquiring SARS-CoV-2 (the virus) and of suffering severe outcomes from COVID-19 (the disease).(2–4) Healthcare workers (HCWs) are also susceptible to infection and may act as vectors, transmitting the virus through necessary daily interactions with patients and amidst imperfect hygiene and infection prevention measures.(1,5) Although the full extent of the ongoing pandemic is unclear and ever-evolving, LTCFs have and continue to bear a disproportionate burden of SARS-CoV-2 infection and COVID-19 mortality.(3,6,7) Across Europe, for instance, LTCFs have accounted for an estimated 30-60% of all COVID-19 deaths as of June 2020.(8)

Effective COVID-19 surveillance is essential for timely outbreak detection and implementation of necessary public health interventions to limit transmission, including case isolation, contact tracing and enhanced infection prevention.(9–1) The current gold-standard diagnostic test for active SARS-CoV-2 infection is RT-PCR (reverse transcriptase polymerase chain reaction), typically performed on clinical specimens from nasopharyngeal swabs.(12) Though sensitive and highly specific, RT-PCR is relatively resource intensive, must be outsourced for institutions lacking on-site infrastructure, and is in many settings subject to shortages and specific usage guidelines. For instance, a common practice in LTCFs in France, the Netherlands, the UK, the USA and elsewhere has been to restrict testing to individuals presenting with characteristic COVID-19 symptoms.(4,13–5) Yet symptomatic infections represent just the tip of the iceberg: many infections cause no or only mild symptoms, produce high quantities of virus in the absence of symptoms, and experience relatively long delays until symptom onset.(16–9) Silent transmission from asymptomatic and pre-symptomatic infections is now a known driver of COVID-19 outbreaks,(20,21) and non-symptomatic cases can act as Trojan Horses, unknowingly introducing the virus into healthcare institutions and triggering nosocomial spread.(8,22,23)

Insufficient surveillance systems, including those lacking testing capacity or relying only on symptoms as indications for testing, have been identified as aggravating factors for COVID-19 outbreaks in LTCFs.(8,16,24–7) Various surveillance strategies have been proposed to optimize testing while accounting for the particular transmission dynamics of SARS-CoV-2, including randomly testing HCWs, testing all patients upon admission, and universal or serial testing.(28–30) Yet COVID-19 surveillance is limited in practice by available testing capacity and health-economic resources, particularly for institutions in low- and middle-income settings.(31,32) In light of testing shortages, group testing (pooling samples from multiple individuals for a single test) has garnered attention as a potentially effective and resource-efficient alternative to standard syndromic surveillance.(33–38)

In order to mitigate and prevent future nosocomial outbreaks, there is an urgent need to optimize COVID-19 surveillance in long-term care settings, taking into account both the unique epidemiological characteristics of SARS-CoV-2 and limited availability of testing resources. Here, we investigated the efficacy, timeliness and resource efficiency of a range of COVID-19 surveillance strategies using simulations from a dynamic SARS-CoV-2 transmission model that uses detailed inter-individual contact data to describe interactions between patients and staff in long-term care.

## METHODS

### Simulating COVID-19 outbreaks in long-term care

We simulated nosocomial outbreaks of COVID-19 using a dynamic, stochastic, individual-based Susceptible Exposed Infectious Recovered (SEIR) model of SARS-CoV-2 transmission coded in C++.(39) The model is described fully in Appendix A using the Overview, Design concepts, and Details (ODD) protocol for individual-based modelling.(40) The goals of our model are to simulate (i) dynamic inter-individual contacts among patients and staff in an LTCF setting, (ii) transmission of SARS-CoV-2 along this simulated contact network, and (iii) clinical progression of COVID-19 among individuals infected with SARS-CoV-2. Throughout, COVID-19 refers to any case of SARS-CoV-2 infection, and not only symptomatic cases.

### Characterizing LTCF structure, demographics, and inter-individual contact behaviour

We used data from the i-Bird study to inform the population structure and dynamic inter-individual contact network used in our model. The i-Bird study has been described elsewhere;(41,42) briefly, close-proximity interactions were measured every 30 seconds by sensors worn by all patients and staff over a 17-week period in a rehabilitation hospital in northern France. There were 170 patient beds across the five wards of this LTCF, and staff were distributed across 13 categories of employment, grouped here as HCWs (caregiver, nurse, physiotherapist, occupational therapist, nurse trainee, physician, and hospital porter) or ancillary staff (hospital services, administration, other rehabilitation staff, management, logistical staff, and activity coordinator/hairdresser) (Figure 1A). This population structure was used in our model, but a novel contact network was simulated to account for missing data resulting from imperfect sensor compliance in the raw contact network (Figure 1B; described in Appendix A). There were on average 170 patients and 240 staff present each week in the simulated network, stratified by ward and type of individual in supplementary Table B1. Contact behaviours were comparable between the raw and simulated networks, and fidelity of the simulated network has been validated previously by its ability to reproduce transmission dynamics from a real outbreak of methicillin-resistant *Staphylococcus aureus* in this LTCF.(41,43)

**Figure 1:**
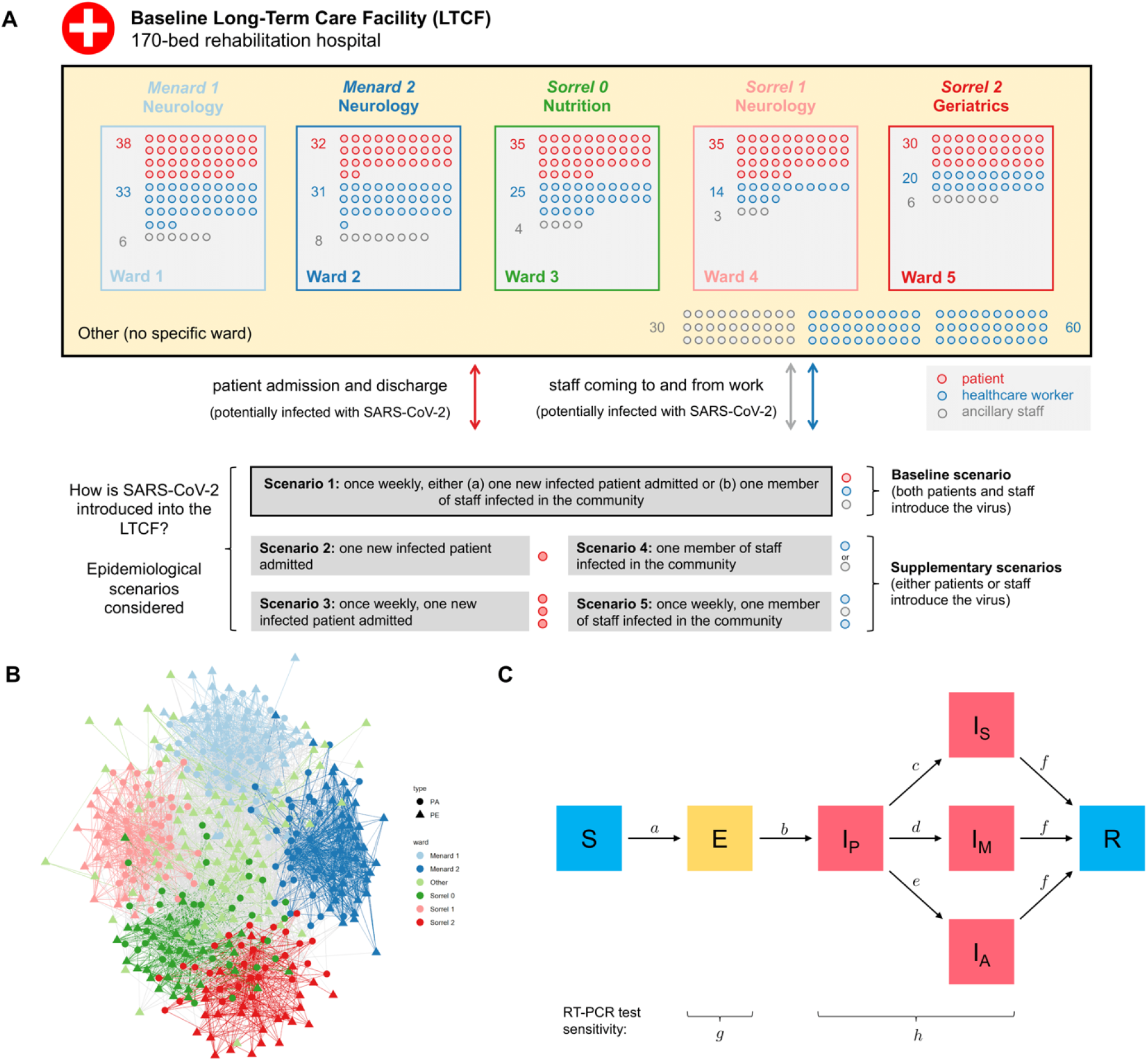
Characteristics of the SARS-CoV-2 transmission model. **A)** A diagram of the baseline LTCF, showing the average weekly number of patients and staff in each ward, including “Other” staff not primarily in any one specific ward. Below the LTCF is a description of the epidemiological scenarios considered for how SARS-CoV-2 was introduced into the LTCF. **B)** A snapshot of the simulated dynamic contact network, showing all patients (PA, circles) and staff (PE, triangles) present in the baseline LTCF as nodes, and inter-individual contacts aggregated over one randomly selected day as edges. Nodes and edges are coloured by ward, with grey edges representing contacts across wards. **C)** A diagram of the modified SEIR process used to characterize COVID-19 infection (S = susceptible, E = exposed, I_P_ = infectious pre-symptomatic, I_A_ = infectious asymptomatic, I_M_ = infectious with mild symptoms, I_S_ = infectious with severe symptoms, R = recovered), with transitions between states *a* to *f* (detailed in supplementary Table A1). We assume low RT-PCR test sensitivity when exposed (*g* = 0.3), and high when infectious (*h* = 0.9).

### Characterizing SARS-CoV-2 transmission and COVID-19 infection

Parameter estimates from the literature were used to characterize SARS-CoV-2 transmission and clinical progression of COVID-19 (see supplementary Table A2). We assumed that susceptible patients and staff could become infected with SARS-CoV-2 if in direct contact with an infectious individual, with the probability of transmission depending on the duration of contact. Assuming *R*_0_=3 for SARS-CoV-2 in the community prior to lockdown,(44) we derived a transmission probability per minute spent in contact *p*=0.14% (see Appendix A).(45)

Clinical progression of COVID-19 was characterized by a modified SEIR process, with: (i) a non-infectious exposed period of 2-5 days, (ii) an infectious pre-symptomatic period of 1-3 days, (iii) an on-average 7-day infectious “symptomatic” period with three levels of symptom severity (severe, mild or asymptomatic), and (iv) eventual recovery with full immunity (Figure 1C). Together, (i) and (ii) amount to an incubation period of 3-8 days, including a 1-3 day window of pre-symptomatic transmission; this is consistent with estimates used elsewhere.(17,19,46) For (iii), we assumed that only 70% of infections ever develop clinical symptoms,(47,48) 20% of which develop severe/critical symptoms.(49) Durations for each stage of infection were drawn probabilistically from uniform distributions for each infection. To reflect rapid clinical deterioration characteristic of severe COVID-19, we assumed no difference in average time to “symptom” onset across asymptomatic, mild symptomatic, and severe symptomatic cases. As surveillance strategies were evaluated only for detection of outbreaks, death and potential long-term clinical outcomes were not explicitly simulated. Each simulation began by introducing one non-symptomatic index case (either exposed, pre-symptomatic or asymptomatic) into the facility on the first day of each simulation (*t*=0). In the baseline scenario, both patients and staff introduced SARS-CoV-2 into the LTCF.

### Measuring outbreaks

Simulated COVID-19 outbreaks were described using various outcome measures, including: infection incidence, infection prevalence, case mix (proportion of infections among patients, HCWs and ancillary staff), outbreak size (cumulative number of cases after 12 weeks of unmitigated transmission), and outbreak size upon first presentation of COVID-19 symptoms.

### Developing a COVID-19 surveillance algorithm

We developed a stochastic surveillance algorithm to evaluate a range of surveillance strategies for their efficacy, timeliness and resource-efficiency in detecting COVID-19 outbreaks. Strategies varied according to who received conventional nasopharyngeal swabs and RT-PCR tests, and with what priority. The algorithm used demographic and clinical indications to administer tests to patients and staff, assuming a daily maximum testing capacity ranging from 1 to 32 tests/day. The algorithm is described in further detail in Appendix A.

### Surveillance strategies considered

Four types of surveillance were evaluated: (i) testing individuals with particular indications, (ii) random testing, (iii) testing cascades, and (iv) group testing (Table 1). Each was further divided into distinct surveillance strategies. For (i), three indications were considered: presentation of severe COVID-like symptoms (reference strategy), presentation of any COVID-like symptoms, or new admission to the LTCF. For (ii), tests were randomly distributed among patients, HCWs, or all patients and staff. In contrast to (i) and (ii), strategies (iii) and (iv) were conceived as hierarchical testing protocols, in which individuals presenting with severe COVID-like symptoms were always tested first to reflect their clinical priority. Remaining tests were subsequently allocated via cascades (iii) or as a single group test (iv).

**Table 1.** Surveillance strategies evaluated for detection of COVID-19 outbreaks in a LTCF. Strategies differ in how swabs and tests are apportioned to patients and staff. Arrows (→) indicate order of priority for testing cascades. Test = RT-PCR test; swab = nasopharyngeal swab; symptoms = COVID-like symptoms; admission = arrival of new patient to the LTCF.

| Surveillance type | Description | Surveillance strategy | Daily testing capacity always reached? |
| --- | --- | --- | --- |
| **Single indication** | Administer tests to any individuals indicated for testing, up to the daily testing capacity. If the number of individuals indicated exceeds the number of tests available, distribute tests randomly among them. | Symptoms (severe) *[reference strategy]* | No |
|  |  | Symptoms (any) | No |
|  |  | Admission | No |
| **Random** | Each day, randomly administer tests to individuals in a particular demographic group. | Random (patients) | Yes |
|  |  | Random (healthcare workers) | Yes |
|  |  | Random (all: patients, healthcare workers and ancillary staff) | Yes |
| **Cascades** | A combination of clinical indications and random testing. First, use indications to administer tests according to a given order of priority. Then, if any tests remain, distribute them randomly among patients not otherwise indicated for testing. | Symptoms (severe) → Symptoms (mild) → Random (patients) | Yes |
|  |  | Symptoms (severe) → Symptoms (mild) → Admission → Random (patients) | Yes |
|  |  | Symptoms (severe) → Admission → Random (patients) | Yes |
|  |  | Symptoms (severe) → Admission → Symptoms (mild) → Random (patients) | Yes |
| **Group testing** | A classical two-stage group testing protocol, modified to account for clinical urgency of severe COVID-19. First, administer individual tests to any patients or staff presenting with severe symptoms. Then, if at least one test remains, pool clinical specimens together and run as one test. If the group test result is positive, individually re-swab and re-test all included individuals. In the baseline, we assume a maximum of 32 swabs per test, but vary this from 2 to 64 swabs per test in sensitivity analysis. | Symptoms (any) | No |
|  |  | Admission | No |
|  |  | Random (patients) (always maximizes number of specimens per group test) | No |
|  |  | Random (HCWs) (always maximizes number of specimens per group test) | No |

For (iii), testing cascades were conceived as mixed testing strategies combining (i) and (ii), in which multiple indications were considered simultaneously but ordered according to their perceived clinical priority. If there were more tests available than individuals indicated for testing, remaining tests were distributed randomly among remaining patients, such that cascades always maximized daily testing capacity. For (iv), clinical specimens from individual swabs were pooled and tested as one ‘group test’, up to a maximum 32 swabs per test in the baseline analysis. SARS-CoV-2 group testing comes at the cost of reduced test sensitivity,(34) which was assumed to decrease linearly with each additional true negative swab (see Appendix A). Various group testing procedures have been proposed elsewhere;(33,35,50-52) here we evaluated a simple two-stage protocol that does not require additional investment or infrastructure, but which requires all individuals included in the initial group test to be re-swabbed and re-tested individually upon a positive group test result in order to determine which individual(s) is (are) infected.(36,53)

### Administering swabs and tests

Each nasopharyngeal swab was coupled with one RT-PCR test, except for group testing strategies, in which multiple swabs were used per test. We assumed a 24-hour lag from swab to test result, perfect test specificity (100%), low test sensitivity (30%) for individuals exposed to SARS-CoV-2, and subsequently high sensitivity (90%) once infectious.(12,54) We further assumed that no individuals refused swabbing/testing. Admission-based tests were administered upon a patient’s arrival to the LTCF, and symptom-based tests were administered on the first day that symptoms appeared. COVID-19 may clinically resemble other acute respiratory infections,(55) such that not all individuals with COVID-like symptoms and indicated for symptom-based testing are actually infected with SARS-CoV-2. Using data from French emergency departments, average daily incidence of influenza-like illness among older adults (50-99 years, 2008-2017) was used as a proxy attack rate for patients and staff presenting with COVID-like symptoms of other aetiologies, 20% of whom were also assumed to present with severe symptoms.(56)

### Surveillance outcomes evaluated

Surveillance strategies were evaluated for their ability to detect COVID-19 outbreaks using four primary outcome measures. First, detection probability, the probability of detecting an outbreak (i) at any time *t* from the index case at *t*=0, (ii) prior to any secondary cases (interpreted as the probability of detecting the index case before any nosocomial transmission), or (iii) prior to first presentation of COVID-19 symptoms. Second, detection lag, the number of days from the index case to outbreak detection (first positive test result). We defined a maximum detection lag of 22 days, after which all outbreaks were assumed to be detected regardless of the surveillance strategy used. Third, outbreak size upon detection, the cumulative number of cases at first positive test result. Fourth, the total number of (i) swabs taken and (ii) tests conducted until outbreak detection; for group testing, this includes resources required to individually re-swab and re-test all individuals included in the initial positive group test.

### Measuring surveillance efficiency

From a health-economic perspective, an efficient use of healthcare resources is one that yields better health outcomes than alternative uses of the same resources.(57) Efficiency can be measured using incremental analysis, in which the additional cost of a particular intervention compared to a reference baseline is scaled by its additional health benefit.(58) This is traditionally expressed as the incremental cost-effectiveness ratio using monetary costs and standardized units of health benefit (e.g., quality-adjusted life-years gained). To report on efficiency in terms of the surveillance cost and benefit outcomes measured in this study, we defined a similar metric, the incremental efficiency ratio (IER),

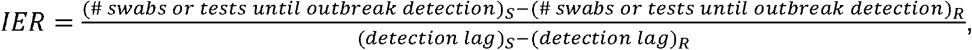

for each surveillance strategy S relative to the reference R. Here, we took the perspective of an LTCF with a reference strategy of only testing individuals with severe COVID-like symptoms. Efficiency results were expressed using the IER as the number of additional swabs and tests required per 1-day improvement in outbreak detection.

### Uncertainty and sensitivity analysis

We ran a range of sensitivity analyses to account for uncertainty in (i) how SARS-CoV-2 was introduced into the LTCF, (ii) the size and structure of the LTCF, and (iii) the transmissibility of SARS-CoV-2 (see assumptions in Appendix A). For each scenario, 100 epidemics were simulated using the transmission model. For each simulated epidemic, the surveillance algorithm was run 100 times across six testing capacities (1, 2, 4, 8, 16 or 32 tests/day), for a total of 60,000 stochastic simulations for each surveillance strategy and each scenario. Across all scenarios, we also varied the maximum number of swabs potentially included per group test (2, 4, 8, 16, 32 or 64 swabs/test). Outcomes were only evaluated for epidemic simulations that resulted in nosocomial outbreaks, defined as simulations with >=1 new case of COVID-19 within 21 days of the initial index case. Unless stated otherwise, outcome measures are reported as median values across all simulations, with uncertainties expressed as 95% uncertainty intervals (UIs), i.e. outcomes from the 2.5th and 97.5th percentiles.

## RESULTS

### SARS-CoV-2 spreads quickly, but COVID-19 symptoms lag behind

SARS-CoV-2 spread quickly, but with a great degree of stochasticity upon its random introduction to simulated LTCFs (Figures 2, B1). After three weeks of unmitigated transmission, a cumulative 86 (95% UI: 6-224) individuals were infected, predominantly other patients (median 72%), then HCWs (25%) and ancillary staff (3%) (Table B2). Outbreaks were characterized by a median lag of 9 (2-24) days between the non-symptomatic index case entering the LTCF and first presentation of mild COVID-19 symptoms among any patient or staff in the facility (Table B3). By the time symptoms emerged, an additional 5 (0-29) individuals had acquired SARS-CoV-2 but were not (yet) showing symptoms (Table B4). Lags were longer for first presentation of severe COVID-19 symptoms (15 days from index case, 4-28), coinciding with a greater cumulative number of secondary infections (25, 0-101).

### Less effective surveillance strategies allow infections to accumulate

Surveillance strategies varied in their ability to detect emerging COVID-19 outbreaks. Surveillance efficacy depended on the stochastic nature of outbreaks, including how many, and which types of individuals became infected over time (Figure 2C,2D). Outbreaks grew exponentially at their outset, so delaying outbreak detection by just one or two days potentially coincided with tens more infections (Figure B2). Five days from the index case entering the LTCF (shortest median detection lag of any strategy), only 2 (1-12) individuals were infected; after ten days, 9 (1-44) were infected; and after fifteen days (longest detection lag at highest testing capacity), 36 (2-124) patients and staff were infected.

**Figure 2.**
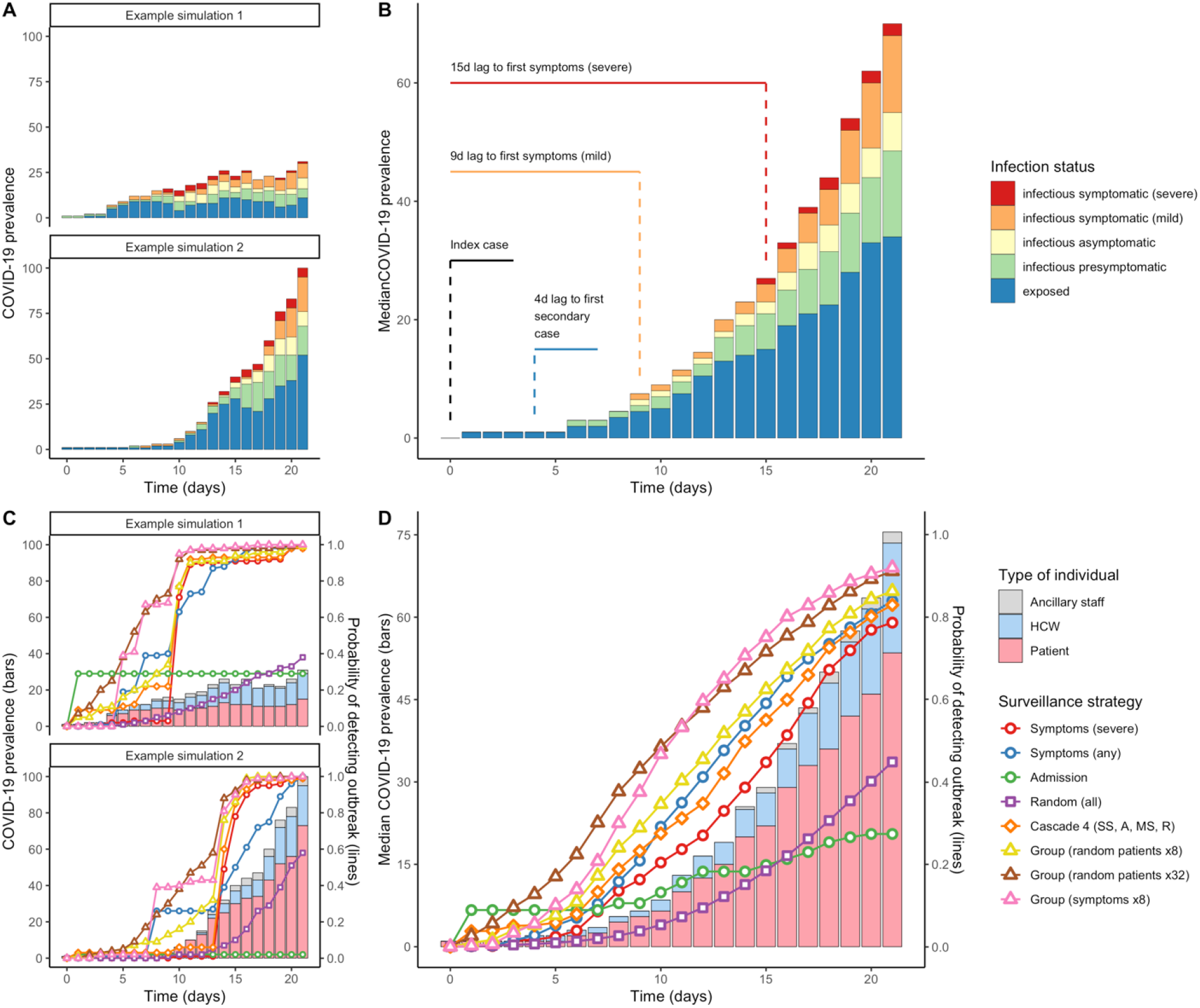
Epidemic curves of COVID-19 infection resulting from random introductions of SARS-CoV-2 into a 170-bed LTCF. Symptomatic cases represent just the “tip of the iceberg” in nascent outbreaks. **(A)** Four examples of epidemic simulations, demonstrating variation in outbreak velocity and lags until first onset of COVID-19 symptoms. **(B)** The median epidemic curve across all simulations for the baseline scenario, with dotted lines demarcating median time lags to selected events. Bars represent the median number of individuals in each infection class over time, and do not necessarily total to the median number infected (e.g. there is a median 1 infection at *t=0* but a median 0 infections in each class, as each index case had an equal 1/3 probability of being exposed, pre-symptomatic or asymptomatic). For the same simulation examples **(C)** and median **(D)**, the probability of detecting outbreaks varied over time for different surveillance strategies (coloured lines), depending on how many, and which types of individuals became infected over time (vertical bars); here, testing capacity = 1 test/day.

### Optimal surveillance depends on daily testing capacity

Across all testing capacities, only testing individuals with severe COVID-like symptoms was among the least effective surveillance strategies considered (Figure 3). This “reference” strategy took a median 15-16 days to detect outbreaks, and had a 3-4% probability of detecting the initial index case prior to any secondary cases (Figure B3). Instead of only severe symptoms, testing individuals with any COVID-like symptoms was more effective, taking 8-14 days to detect outbreaks, with a 4-16% probability of detecting index cases prior to any secondary cases. Only testing patients at admission was overall ineffective by right of detecting neither staff index cases nor ongoing outbreaks already underway in the LTCF, resulting in long median delays to outbreak detection (11-22 days) despite comparatively high probabilities of detecting COVID-19 prior to any secondary cases (9-35%). (In the scenario where only new patients introduced SARS-CoV-2 into the LTCF, when screening all admissions there was a 66% probability of detecting the index case upon admission and hence prior to transmission.) For random testing strategies, surveillance was highly ineffective when few tests were available, but increasingly effective at higher testing capacities. Conversely, for indication-based strategies, efficacy plateaued when capacity exceeded the number of individuals indicated for testing (Figure B3).

**Figure 3.**
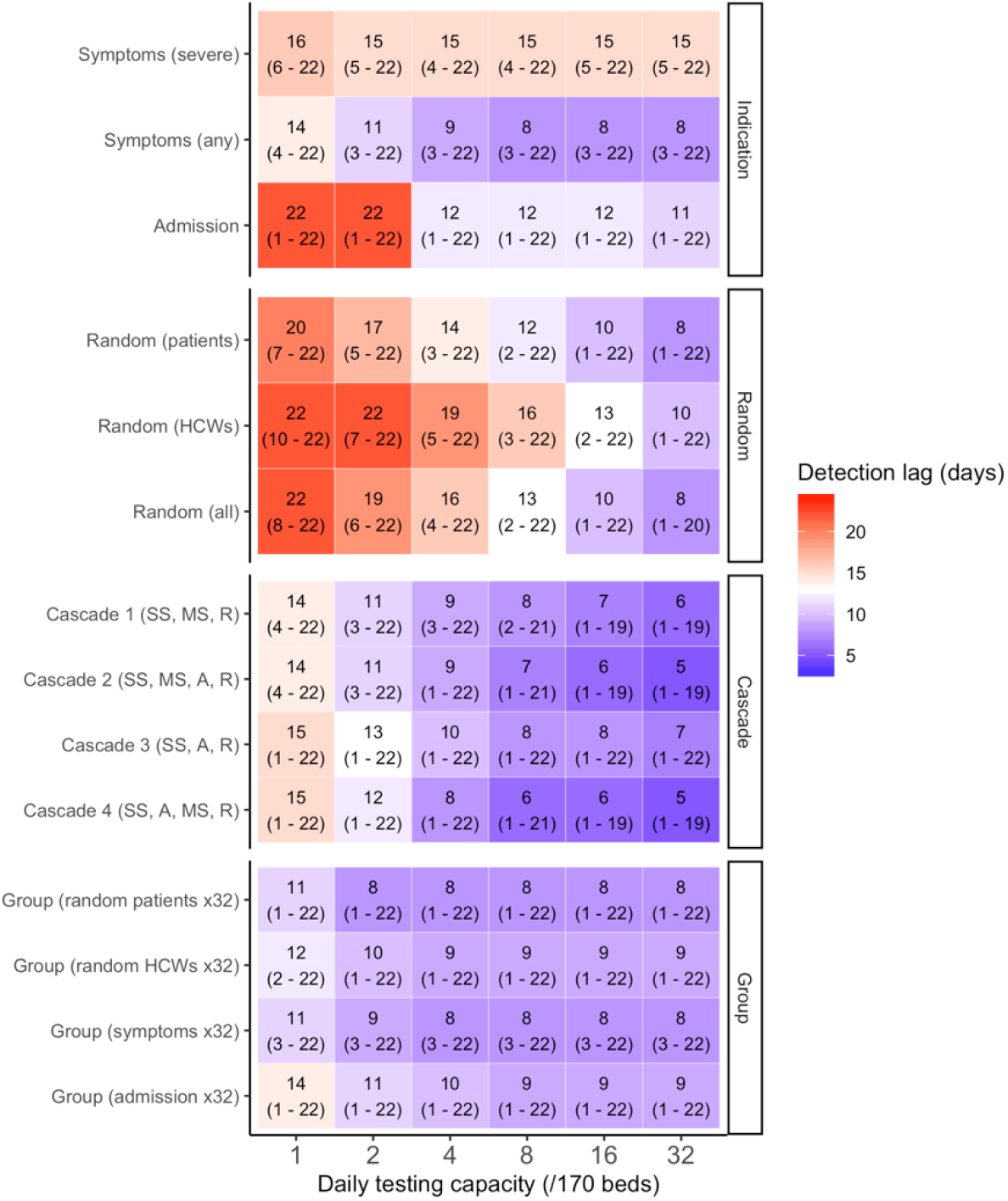
Test more to detect outbreaks sooner. Median lags to outbreak detection (95% uncertainty interval) are shown for each surveillance strategy (y-axis) as a function of the daily testing capacity (x-axis). Group testing strategies assume a maximum of 32 swabs per test. For both cascades and group testing, individual tests were always reserved for individuals with severe COVID-like symptoms; remaining tests were then distributed according to cascades or as a single group test. SS = severe symptoms; MS = mild symptoms; A = admission; R = random patients.

At high testing capacity (16-32 tests/day, ≈1 test/5-11 beds/day), testing cascades were the most effective surveillance strategies. The four cascades considered here detected outbreaks within a median 5-8 days, coinciding with just 1-4 COVID-19 infections among all patients and staff. Cascades had a 23-52% chance of detecting outbreaks prior to any secondary cases, a 38-63% chance prior to the emergence of any COVID-19 symptoms, and an 84-91% chance prior to severe COVID-19 symptoms. Cascades that included both new patient admission and presentation of any COVID-like symptoms as indications for testing were most effective.

At low testing capacity (1 or 2 tests/day, ≈1 test/85-170 beds/day), group testing was the most effective form of surveillance considered. Compared to the reference (16 days) and cascades (14-15 days), outbreaks were detected within 8-11 days (coinciding with a cumulative 5-9 infections) when pooling random patients, or 10-12 days (9-15 infections) when pooling random HCWs. At this low capacity, it was also more effective to pool symptomatic individuals in group tests (9-11 days, 7-13 infections) than to test them individually (11-14 days, 13-21 infections) because individuals with non-COVID but COVID-like symptoms were also “in competition” for limited tests. Compared to the baseline protocol, which assumed a maximum of 32 swabs/test, group testing was less effective given fewer swabs per test, despite potentially higher test sensitivity. For example, when pooling randomly selected patients in daily group tests, outbreaks were detected within 8-11 days at 32 swabs/test, 10-13 days at 8-16 swabs/test, and 11-16 days at 2-8 swabs/test (Figure B4).

### Group testing symptomatic individuals is the most efficient use of both swabs and tests

Surveillance strategies varied considerably in their use of testing resources (Figure B5) and in their efficiency for improving COVID-19 outbreak detection relative to the reference strategy (Figure 4). The reference used the fewest swabs and tests, on average <1/day regardless of the assumed daily testing capacity (owing to a low daily incidence of severe COVID-like symptoms). At high testing capacity (16-32 tests/day), the high incremental efficacy of cascades (outbreak detection 7-10 days earlier than the reference, prior to 19-22 additional infections) resulted from extensive resource use (83-211 additional tests and swabs), for efficiency ratios of IER=9.2-26.4 additional swabs and tests per 1-day improvement in outbreak detection. Simply allocating individual tests to patients and staff with any COVID-like symptoms was a more efficient means to improve surveillance (detection=7 days earlier, IER=3.4 additional swabs and tests / 1-day improvement in outbreak detection).

**Figure 4.**
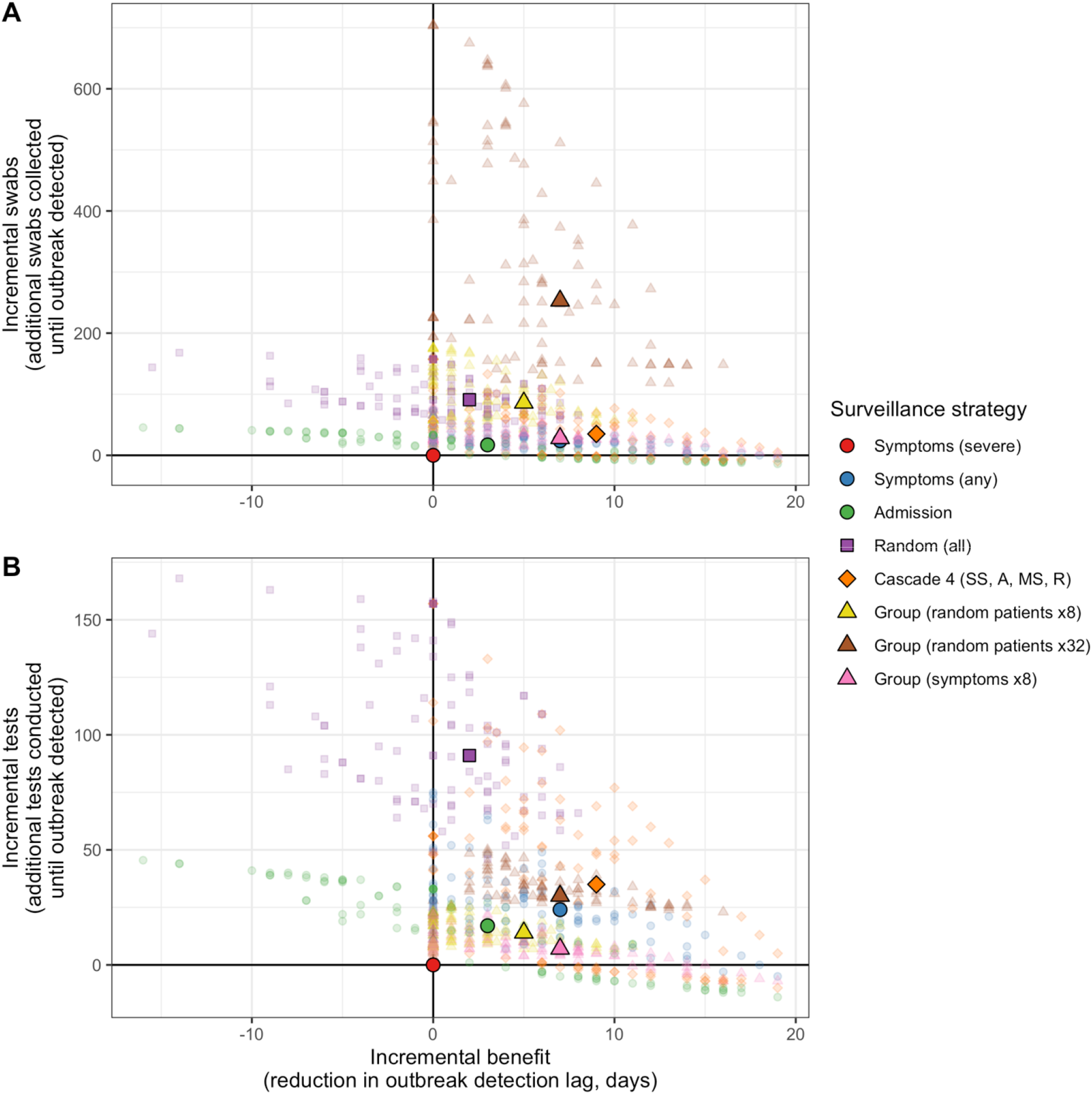
Incremental efficiency plots for selected surveillance strategies relative to a reference strategy of only testing individuals with severe COVID-like symptoms, balancing improvement in COVID-19 surveillance (x-axis) against additional nasopharyngeal swabs used (y-axis for **A**) and additional RT-PCR tests conducted (y-axis for **B**) until outbreaks were detected. For both plots, daily testing capacity is fixed at 8 tests/day, small translucent points represent median outcomes across all surveillance simulations for each simulated outbreak, and larger opaque points represent median outcomes across all outbreaks.

Group testing strategies were generally efficient with respect to tests, but used highly variable numbers of swabs to detect outbreaks. At high swabbing capacity (16-32 swabs/group test, ≈1 swab/5-11 beds/day), pooling randomly selected patients used a median 18-30 excess tests to detect outbreaks 4-7 days earlier (IER_tests_=3.0-5.6), but a median 80-254 additional swabs (IER_swabs_=20.0-36.3). Results were similar when pooling randomly selected HCWs (detection=5-6 days earlier, IER_tests_=3.2-4.5, IER_swabs_=32.2-47.5). By contrast, for all scenarios considered, pooling individuals with any COVID-like symptoms was among the most efficient strategies in terms of both swabs and tests. In the most resource-limited scenarios (1-2 tests, ≈1 test/85-170 beds/day; 2-8 swabs per group test, ≈1 swab/21-85 beds/day), it was both the most effective means to detect COVID-19 outbreaks (detection=3-6 days earlier, prior to 10-18 additional infections) and the most efficient means to improve surveillance from the reference (4-7 additional tests, 9-24 additional swabs; IER_tests_=0.8-1.4, IER_swabs_=2.8-4.0).

## DISCUSSION

The ongoing COVID-19 pandemic continues to devastate LTCF populations worldwide, with high rates of mortality among particularly frail and elderly patients, and high rates of infection among patients and staff alike.(3,5,6,8) This motivates a need for timely and efficient surveillance strategies that optimize limited testing resources to detect outbreaks as quickly as possible. We used an individual-based transmission model to simulate COVID-19 outbreaks in LTCF settings, and compared a range of strategies for how to allocate limited nasopharyngeal swabs and RT-PCR tests to detect these outbreaks as quickly and efficiently as possible, under a range of epidemiological scenarios.

Our findings suggest that LTCFs with ample testing resources can detect emerging COVID-19 outbreaks most quickly by using testing cascades. The most effective cascades considered multiple indications, including both COVID-like symptoms and patient admission, and detected outbreaks days ahead of traditional symptom-based screening. By extension, of the strategies considered here, cascades had the greatest probability of identifying non-symptomatic cases. These findings held in sensitivity analyses considering outbreaks in a smaller, 30-bed geriatric LTCF (Figure B6), as well as when assuming unusually low or high SARS-CoV-2 transmission rates (Figures B7, B8). Although only a select few indications were considered in the present study, LTCFs may consider a wider range of known risk factors for SARS-CoV-2 acquisition in their own cascades to maximize the probability of detecting emerging outbreaks before widespread transmission.

COVID-19 surveillance was less effective in resource-limited settings because of an inability to regularly test large numbers of patients and staff. However, resource limitations were in part overcome by group testing: in our analysis, group testing was the most effective means of COVID-19 surveillance under limited testing capacity, and group testing strategies were among the most resource efficient means to improve surveillance outcomes with respect to a "bare minimum” reference of only testing individuals with severe COVID-like symptoms. Recent mathematical modeling results have further suggested that group testing could be cost-effective for COVID-19 screening in large populations.(33) As with cascades, LTCFs interested in group testing may consider a wider range of indications than was possible to include in this study, in order to maximize the probability of including potentially infected patients and staff in routine group tests.

Our analysis was limited to classical two-stage group testing, initially proposed by Dorfman in 1943 for syphilis screening among World War II soldiers,(53) in which all individuals in a positive group test are individually re-tested to determine who is infected. This is regarded as the most straightforward approach,(36) and we conservatively assumed re-swabbing in addition to re-testing of all individuals in a positive group test to account for potential logistical challenges of storing and maintaining large numbers of swabs for re-testing. Various alternative group testing strategies have been proposed and implemented elsewhere, including the use of simultaneous multi-pool samples, non-adaptive pooling schemes, and others.(36,38,51,52) These have the advantage of not requiring separate re-testing of all individuals in a positive group test, and are hence more efficient in terms of the number of tests required for case identification. However, these strategies may also require additional testing infrastructure and expertise, which may be cost-prohibitive for the resource-limited settings that may benefit most from group testing in the first place. Decision-makers must consider trade-offs between the various costs and benefits of different group testing technologies, including how many individuals to include per test, and how many stages of testing to conduct.

We predicted that silent introductions of SARS-CoV-2 led to large outbreaks in the absence of specific control strategies. This is consistent with large COVID-19 outbreaks observed in LTCFs worldwide,(6,8,10,16) including an infamous outbreak in King County, Washington that resulted in 167 confirmed infections within three weeks of the first reported case.(5) We further predicted that larger proportions of patients became infected than staff, consistent with emerging evidence of higher SARS-CoV-2 incidence in patients than staff across LTCF settings worldwide.(3,8) We also predicted larger and more rapid outbreaks when SARS-CoV-2 was introduced through admission of an infected patient, rather than through a member of staff infected in the community, with important implications for surveillance efficacy (Figure B9). These findings are likely due to the nature of human interactions in the LTCF upon which we based our model, in which patient-patient contacts were particularly long and numerous.(42) Overall, these findings reinforce both (i) a need to screen incoming patients potentially exposed to or infected with SARS-CoV-2,(59) and (ii) the importance of interventions to limit contact between patients (e.g. isolation of retirement home residents), as already widely recommended for affected facilities in the current pandemic context.(4)

Simulated outbreaks were further characterized by delays between silent introduction of SARS-CoV-2 and first onset of COVID-19 symptoms, during which time new infections not (yet) showing symptoms accumulated. This is consistent with reported transmission dynamics of SARS-CoV-2; for instance, modelling studies have estimated that 37-48% of secondary infections among hospital transmission-pairs resulted from pre-symptomatic transmission,(60) and that, early on in COVID-19 outbreaks, transmission events occur on average two to three days prior to symptom onset.(17) Findings are also consistent with high proportions of asymptomatic infection, and important roles for pre-symptomatic and asymptomatic transmission reported in various LTCF outbreaks.(5,16,21,26–29) The often silent nature of SARS-CoV-2 transmission highlights epidemiological challenges associated with screening for emerging outbreaks using symptoms alone. In addition to the strategies highlighted above, we found that testing patients and healthcare workers with any, and not only severe COVID-like symptoms can substantially improve outbreak detection, supporting recommendations to expand testing criteria in LTCFs to include individuals with atypical signs and symptoms of COVID-19, such as muscle aches, sore throat and chest pain.(59)

A strength of the present study is the use of highly detailed inter-individual contact data to inform our individual-based transmission model. This allowed for recreation of life-like interaction dynamics among and between LTCF patients and staff, and facilitated simulation of more realistic SARS-CoV-2 dissemination than a traditional mass-action transmission process. We are aware of no other studies using detailed individual-level contact networks to simulate SARS-CoV-2 transmission, nor of studies using transmission modelling to evaluate COVID-19 surveillance strategies for healthcare settings.

This work has several limitations. First, substantial uncertainties remain regarding epidemiological characteristics of COVID-19, including the exact proportion of asymptomatic infections in LTCFs,(23,27,61,62) the transmissibility of SARS-CoV-2 over the course of infection and per unit of time spent in contact, and a potential role for environmental acquisition,(63–65) which was not included in our model. Furthermore, many LTCFs have already implemented control measures, such as interruption of social activities and provisioning of personal protective equipment, that should act to reduce transmission from baseline. We conducted sensitivity analyses to consider unusually high and low transmission rates to reflect these uncertainties. Although SARS-CoV-2 spread more or less quickly, the relative efficacies of surveillance strategies were largely unchanged in these analyses, resulting in the same conclusions for optimizing use of limited testing resources to detect COVID-19 outbreaks (Figures B7, B8, B10, B11).

Second, LTCFs represent a diverse range of healthcare institutions, each with unique specializations, patient populations and living conditions, and the generalizability of our findings across these settings is not clear. For this reason, simulations restricted to the geriatric ward were conducted to approximate a smaller LTCF geared towards elder care. In this ward, simulated contact patterns (8.0 daily patient-patient contacts, 8.3 daily patient-staff contacts) were comparable to patterns observed in a nursing home in Paris (5.0 daily patient-patient contacts, 6.3 daily patient-staff contacts),(66) and corresponding results may be interpreted as what may occur in a nursing home environment. In this 30-bed LTCF, high testing and swabbing capacities approximated universal testing strategies, in which large proportions of individuals were routinely tested. This explains why randomly testing among all individuals was the single most effective strategy at highest testing capacity (Figure B6), and why pooling randomly selected individuals was a particularly effective strategy in this setting (Figure B12). Otherwise, conclusions for surveillance were similar to the baseline LTCF.

Finally, the testing landscape for COVID-19 is due to shift quickly, with increased testing capacity and alternative testing technologies, such as rapid diagnostic tests, likely to become increasingly available in the coming months and years. However, uptake of new technologies is certain to be heterogeneous, and testing resources may remain limited for the foreseeable future, particularly in low- and middle-income settings.(31,32) Although we explicitly modelled standard RT-PCR testing, our findings may be broadly generalizable to other COVID-19 testing technologies with limited capacity. Findings for group testing, however, necessarily assume that pooling samples from multiple individuals is both logistically feasible and retains sufficient test sensitivity, as demonstrated for RT-PCR. Further, even in settings with abundant testing capacity, limiting the number of tests necessary to detect an outbreak will remain a priority given health-economic concerns.

## Conclusions

In conclusion, our findings demonstrate the susceptibility of LTCFs to large COVID-19 outbreaks, and suggest that testing cascades, which combine clinical indications and random testing, are a highly effective means to detect emerging outbreaks given ample testing resources. For resource-limited settings unable to routinely screen large numbers of individuals, however, group testing is a preferable strategy, both more effective and resource-efficient than cascades and other considered strategies. These findings add to a limited evidence base for optimizing COVID-19 surveillance in healthcare institutions. Even in regions where non-pharmaceutical interventions have come to slow transmission in the community, LTCFs remain uniquely vulnerable to COVID-19. Increasing testing capacity and expanding surveillance beyond symptom-based screening could allow for earlier outbreak detection, facilitating timely intervention to limit transmission and save lives.

## Data Availability

N/A

## Ethics approval and consent to participate

This study obtained all authorizations in accordance with French regulations regarding medical research and information processing. All French IRB-equivalent agencies accorded the i-Bird program official approval (CPP 08061; Afssaps 2008-A01284-51; CCTIRS 08.533; CNIL AT/YPA/SV/SN/GDP/AR091118 N°909036). Signed consent by patients and staff was not required according to the French Ethics Committee to which the project was submitted.

## Availability of data and materials

The datasets used and/or analysed during the current study are available from the corresponding author on reasonable request.

## Authors’ contributions

LO and LT conceived the study. AD programmed the transmission model. DS programmed the surveillance algorithm, analyzed the data and presented results. DS, AD, LO and LT interpreted the results. KP, BTH, DG and JF consulted on analyses. DS wrote the article. All authors edited and revised the article.

## Acknowledgements

This work resulted from the MOD-COV Project (Modelling of the hOspital Dissemination of SARS-CoV-2), a collaboration between the Institut Pasteur, the Conservatoire Nationale des Arts et Métiers, and the AVIESAN/REACTing working group “Modelling SARS-CoV-2 dissemination in healthcare settings”, whose members we thank (Niccolo Buetti, Christian Brun-Buisson, Sylvie Burban, Simon Cauchemez, Guillaume Chelius, Anthony Cousien, Pascal Crepey, Vittoria Colizza, Christel Daniel, Aurélien Dinh, Pierre Frange, Eric Fleury, Antoine Fraboulet, Marie-Paule Gustin, Lidia Kardas-Sloma, Elsa Kermorvant, Jean Christophe Lucet, Chiara Poletto, Rodolphe Thiebaut, Sylvie van der Werf, Philippe Vanhems, Linda Wittkop, Jean-Ralph Zahar).

We also thank the i-Bird Study Group (Anne Sophie Alvarez, Audrey Baraffe, Mariano Beiró, Inga Bertucci, Pierre-Yves Boëlle, Camille Cyncynatus, Florence Dannet, Marie Laure Delaby, Pierre Denys, Matthieu Domenech de Cellès, Eric Fleury, Antoine Fraboulet, Jean-Louis Gaillard, Boris Labrador, Jennifer Lasley, Christine Lawrence, Judith Legrand, Odile Le Minor, Caroline Ligier, Lucie Martinet, Karine Mignon, Catherine Sacleux, Jérôme Salomon, Thomas Obadia, Marie Perard, Laure Petit, Laeticia Remy, Anne Thiebaut, Damien Thomas, Philippe Tronchet, Isabelle Villain), as well as Jacob Barrett for helpful discussion.

## **Appendix A:** Supplementary methods for the article *Optimizing COVID-19 surveillance in long-term care: a modelling study*

### Introducing a SARS-CoV-2 transmission model for long-term care settings

Nosocomial COVID-19 outbreaks were simulated using a dynamic, stochastic, individual-based model (IBM) coded in C++ with three main goals: (i) to use detailed inter-individual contact data to simulate dynamic contact networks among patients and staff in long-term care facility (LTCF) settings, (ii) to simulate transmission of SARS-CoV-2 among LTCF patients and staff in simulated contact networks, and (iii) to simulate clinical progression of COVID-19 among individuals infected with SARS-CoV-2 using a Susceptible Exposed Infectious Recovered (SEIR) process (throughout, the virus is referred to as SARS-CoV-2, and human infection with this virus as COVID-19). We subsequently developed a surveillance algorithm to evaluate different surveillance strategies for detection of simulated COVID-19 outbreaks.

First, we broadly introduce the data used to inform and parameterize the transmission model. Second, we describe the transmission model in full using the Overview, Design concepts, and Details (ODD) protocol for individual-based modelling. Third, we describe the surveillance algorithm and evaluation of different surveillance strategies.

### I. Data informing the individual-based model

#### Characterizing dynamic inter-individual contacts in an LTCF setting

As described in the main text, LTCF structure, demographics, and dynamic contact networks were estimated using data from the i-Bird study. A statistical analysis of the i-Bird contact network has been published previously.(1) Briefly, using nearly 2.7 million close-proximity interactions (CPIs) recorded from 318 patients and 262 staff between July and October 2009, distinct contact patterns were identified for individuals in each ward, reflecting behaviours particular to patients and different types of staff in this LTCF. For instance, patients typically spent 24 hours per day in the facility and had higher rates of contact with HCWs during mornings and afternoons, but with other patients in evenings. Patients were potentially admitted or discharged over the course of the study period. The average patient length of stay was 7 weeks and a median 2 (range 0 - 11) new patients were admitted per day. Staff were present according to their respective working hours, and had fewer overall contacts during evenings and weekends. HCWs had more distinct contacts with other individuals (on average 14.3/day) than patients (11.2/day), but had a shorter cumulative duration of time spent in contact with others (15 minutes/day) than patients (32 minutes/day). Compared to other wards, contacts were fewest (8.6 distinct CPIs/day) and longest (47 cumulative minutes/day) in the geriatric ward. Further, in contrast to a contact network observed using similar methods in an acute care setting, patient-patient CPIs were particularly frequent and numerous in this LTCF.(1,2)

#### Characterizing SARS-CoV-2 transmission along human contact networks

In the transmission model, Susceptible patients and staff could become infected with COVID-19 if in direct contact with an Infectious individual. We assumed that the probability of transmission per infectious contact depends on the duration of that contact, *d* (limited to intervals of 30 seconds, the discrete time-step for the transmission model). This transmission probability was computed as follows. Assuming homogeneous mixing among individuals, the basic reproduction number (*R*_0_) of a pathogen can be approximated as

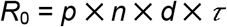

where *p* is the per-minute probability of transmission between Susceptible and Infectious individuals in contact with one another; *n* is the average number of daily contacts per individual; *d* is the average duration of these contacts; and *τ* is the duration of the infectious period.(3) Using community estimates from France, including epidemiological modelling of COVID-19 transmission prior to lockdown (*R*_0_=3)(4) and a detailed survey of inter-individual contacts in the general community (*n* = 8 contacts/days, d = 30 minutes),(5) and assuming an infectious period of *τ* = 9 days for COVID-19, we calculated a transmission probability of *p* = 0.14% per minute spent in contact with a Susceptible individual. We further set a saturation threshold at one hour of contact, such that the per-contact transmission probability was at most 8.3% per contact between any two individuals. To reflect uncertainty in the transmissibility of COVID-19 per infectious contact, in sensitivity analyses we considered extreme estimates for COVID-19 epidemicity in the community (*R*_0_=1.5, 6), which translate to low and high transmission probabilities per minute spent in contact (*p*=0.07%, 0.21%).

#### Characterizing COVID-19 infection

The natural history of COVID-19 infection was conceptualized as a modified SEIR process. Corresponding with the model illustration in Figure 1C of the main text, the transitions of this model are described below in Table S1.

**Table A1.** State transitions for the SEIR infection process. Numerical values are drawn probabilistically for each transition for each individual in the IBM.

| Symbol | Name | Transition | Description |
| --- | --- | --- | --- |
| a | Infection | $S \rightarrow E$ | After a contact with an Infectious individual, Susceptible individuals become infected and enter Exposed class with probability: $(p \times \text{duration of contact})$ |
| b | Infectiousness onset | $E \rightarrow I_P$ | Exposed individuals become Infectious (but do not yet show symptoms) at rate: $(1/d_E)$ |
| c | Symptom onset (severe) | $I_P \rightarrow I_S$ | Pre-symptomatic Infectious individuals develop severe symptoms at rate $(1/d_P) \times (s_{MS}) \times (s_S)$ |
| d | Symptom onset (mild) | $I_P \rightarrow I_M$ | Pre-symptomatic Infectious individuals develop mild symptoms at rate $(1/d_P) \times ((s_{MS}) - (s_{MS} \times s_S))$ |
| e | Symptom onset (asymptomatic) | $I_P \rightarrow I_A$ | Pre-symptomatic Infectious individuals progress to the asymptomatic Infectious state at rate $(1/d_P) \times (1 - s_{MS})$ |
| f | Recovery | $(I_S \text{ or } I_M \text{ or } I_A) \rightarrow R$ | Individuals progress to a Recovered state (non-infectious, non-symptomatic, non-susceptible to re-infection) at rate $(1/d_S)$ |

As described in the main text, clinical progression of COVID-19 was characterized by: (i) a non-infectious exposed period of 2-5 days, (ii) an infectious pre-symptomatic period of 1-3 days, (iii) an on-average 7-day infectious “symptomatic” period with three levels of symptom severity (severe, mild or asymptomatic), and (iv) eventual recovery with full immunity.

Similar infection processes have been used in contemporaneous COVID-19 modelling studies,(4,6–8), and sources for assumed parameter values are provided in parameter Table S2. Here, this structure allows for (i) a 3-8 day incubation period, (ii) 1 to 3 days of pre-symptomatic transmission, as well as (iii) potential asymptomatic transmission in individuals who never show symptoms.

**Table A2.** Model parameter estimates.

| Parameter | Value (distribution) | Source |
| --- | --- | --- |
| **Parameters for estimation of SARS-CoV-2 transmission rate per minute of contact** |  |  |
| SARS-CoV-2 basic reproduction number ($R_0$) in the general community prior to lockdown | 3.0 (1.5, 6.0 in sensitivity analysis) | (4) |
| Average infectious period of SARS-CoV-2 ($\tau$) | 9 days | (9) |
| Average number of contacts per day per individual prior to lockdown ($n$) | 8 | (5) |
| Average duration per contact prior to lockdown ($d$) | 30 minutes | (5) |
| **Epidemiological and clinical parameters** |  |  |
| SARS-CoV-2 transmission rate per minute of contact ($p$) | 0.00139 (0.00070, 0.00278 in sensitivity analysis) | Estimated |
| Duration of exposed period (latency) ($d_E$) | 2-5 days (uniform) | Approximated from (10,11) |
| Duration of pre-symptomatic period ($d_P$) | 1-3 days (uniform) | Approximated from (10,11) |
| Duration of symptomatic period (whether asymptomatic, mild symptomatic or severe symptomatic) ($d_S$) | 7 days (log-normal, $\sigma^2 = 7$) | Approximated from (12) |
| Proportion of COVID-19 infections presenting any symptoms ($s_{MS}$) | 0.7 | Approximated from (13,14) |
| Proportion of symptomatic COVID-19 infections with severe symptoms ($s_S$) | 0.2 | (15) |
| **Testing and surveillance parameters** |  |  |
| Daily incidence proportion of non-COVID but COVID-like symptoms | 0.011 | Estimated from OSCOUR data, described in (16) |
| Proportion of non-COVID but COVID-like symptoms with signs of severity | 0.2 | Assumed |
| Delay from test to test result | 1 day | Assumed |
| Test specificity | 100% | (17) |
| Test sensitivity (infectious stages of infection) | 90% | Approximated from (17,18) |
| Test sensitivity (exposed stage of infection) | 30% | Approximated from (17,19) |
| Daily testing capacity (tests/day) | 1, 2, 4, 8, 16, 32 | Assumed |
| Maximum number of specimens per group test | 2, 4, 8, 16, 32, 64 | Assumed |
| RT-PCR sensitivity discounting rate per additional true-negative specimen | 0.3125% | Estimated from (20) |

### II. Description of the model using the ODD protocol

The simulation model used in this study is CTCModeler, an individual-based model (IBM) developed to simulate pathogen transmission among and between patients and staff in healthcare settings.(21) The IBM was programmed in C++ with the repast HPC library 2.2.0. In the following, we describe the IBM as it was used for the present study, following the ODD protocol of Grimm et al.(22)

#### 1. Purpose

The IBM simulates nosocomial transmission of SARS-CoV-2 between humans using inter-individual contact data. The main goals are (i) to simulate inter-individual contacts between individuals in an LTCF, (ii) to trace SARS-CoV-2 transmission along this contact network, and (iii) to describe clinical progression of COVID-19 infection among individuals infected with SARS-CoV-2.

#### 2. Entities, State Variables and Scales

##### Entities and state variables

All entities in the model are conceived as existing within the LTCF. There are two main classes in the model: Individual and Pathogen. These are derived from the abstract Organism class, which is composed of common variables inherited by both Individual and Pathogen. The Individual class has two children, patient and staff (visitors were excluded). Pathogen has only one child, the virus SARS-CoV-2. Each object has a unique ID variable in each simulation.

Common variables for Individuals are anonymous hospital ID number, age, gender, type of individual (patient or staff), admission date, discharge date, allocated ward, a map of infection, current positive SAR-CoV-2 infection, and infection status (statuses following a modified SEIR process are described above). For patients, admission refers to arriving to the LTCF as a new patient, and discharge refers to leaving the LTCF; for staff, admission refers to coming to work, and discharge refers to leaving work.

The map of pathogen status contains the current status of infection as well as its duration, i.e. the date at which an individual passes from one infection state to the next. The patient class also includes a hospital flag variable, which describes the reason for hospitalization, while the staff class also includes a category variable, which describes that individual’s occupation. There are 13 occupations in total: caregiver, nurse, physiotherapist, occupational therapist, nurse trainee, physician, hospital porter, hospital services, administration, other rehabilitation staff, management, logistical staff, and activity coordinator/hairdresser.

The common variable for Pathogen is current positive individuals, which lists all individuals who are infected with the virus at a given point in time.

##### Scales

The model runs using discrete 30-second time-steps and each simulation is run for approximately 12 weeks (85 days).

There is no explicit spatial scale in the model; instead, individuals enter into contact with one another according to hourly probabilities of contact, stratified by type of individual, occupation (for staff), ward, time of day and day of the week (weekday vs. weekend). Simulation of this contact network is detailed further below.

#### 3. Process Overview and Scheduling

Simulation processes are run on three time schedules: weeks, days and time steps.

##### Weeks

Each week, a spontaneous infection can occur either in a new patient upon their admission to the LTCF, or among admitted staff. The latter is conceptualized as infection occurring in the community (i.e. outside the LTCF). This scheduling process is only relevant for epidemiological scenarios with weekly introductions of SARS-CoV-2 (scenarios 1, 3, and 5). See the Details (initialization) section for more information on epidemiological scenarios considered.

##### Days

The model performs three actions on Individuals each day: change infection status, add individual (admission), or remove individual (discharge). Infection progression follows an SEIR process, described above and in the main text. Durations for each stage of infection for each individual are drawn probabilistically from uniform distributions (parameter values in Table A1). Individuals are added or removed based on daily admissions and discharges listed in the admission file from the i-Bird study. Readmissions are possible for both patients and staff. When individuals "leave” the simulation, they go into a transitory container that keeps track of their information (e.g. immunization status) in case later re-admitted.

##### Time Step

The model time step is 30 seconds. At each time step, the model simulates (i) contacts and (ii) possible SARS-CoV-2 transmission events between individuals, which are calculated using the sum total of the number of time steps in each contact.

#### 4. Design concepts

##### Basic principles

This IBM uses detailed inter-individual contact data from the i-Bird study to build a stochastic, dynamic contact network between all Individuals in the LTCF over a 12-week period. Individuals with a ‘Susceptible’ infection status can become infected with SARS-CoV-2 if in contact with an Individual with an ‘Infectious’ infection status. This infection process is based on classical concepts from the mathematical modelling of infectious diseases. Both of these processes are conceptualized as ‘Interactions’ and are described below.

##### Interactions

Two types of interactions occur during the simulation:

#### Inter-individual Contacts

Patient and staff populations of the Berck-sur-Mer rehabilitation hospital in Northern France are used in the model. This includes 318 patients and 262 staff included in the i-Bird study and present in the LTCF over a 12-week period, described elsewhere.(1) Patients and staff are distributed across five wards: Menard 1 (neurology), Menard 2 (neurology), Sorrel 0 (nutrition), Sorrel 1 (neurology), and Sorrel 2 (geriatrics). 90 members of staff are not affiliated with one particular ward and are instead grouped as being in the ‘Other’ ward. This includes both staff working in back offices (i.e. not on any particular ward), as well as staff who regularly work across multiple wards.

Real staff scheduling and patient admission data are used to determine who is ‘admitted’ in the model (i.e. present in the LTCF) on any given day of the 12-week simulation period. However, instead of using the raw contact measured in the i-Bird study, a novel contact network is simulated to account for missing data resulting from imperfect sensor compliance in the raw network.

For the simulated network, contact probabilities are estimated using hourly contact rates from the raw network for each type of individual, ward, time of day, and day of the week (weekday vs. weekend). Contact durations are drawn from log-normal distributions; duration means and standard deviations also estimated from the raw network.

At each time step, the model builds an edge between pairs of Individuals if newly in contact with one another (based on contact probabilities described above), maintains the edge if still in contact (based on contact durations described above), and removes the edge once no longer in contact.

In a previous analysis, the simulated network was found to have a greater total number of contacts than the raw network, as expected to account for missing data, but contact durations were nearly identical and other network properties were similar; for instance, the degree of the patient-to-staff subgraph for the simulated network (8.98, 8.85 – 9.12) is comparable to the raw network (8.71, 8.52 – 8.91).(23)

#### Inter-individual Transmission

Infectious Individuals can transmit their Pathogen to Susceptible Individuals through the contact network. For each contact, the probability of transmission per minute spent in contact (*p*) rate is multiplied by the duration of the contact in minutes (*d*). Here, the model also incorporates transmission probability saturation at 60 minutes (such that *p* <= 8.3%). The infection process has no bearing on the infection status of the Individual acting as the Pathogen donor.

##### Stochasticity

For each model scenario, CTCModeler is run 100 times to produce 100 distinct epidemic simulations (see details of different model scenarios below). This is done to account for stochasticity in three model process: (i) initialization conditions, (ii) COVID-19 life history, and (iii) SARS-CoV-2 transmission.

For (i), the first Individual infected with SARS-CoV-2 at t=0 (the index case) is randomly selected among patients newly admitted that same day, or among staff presently admitted (depending on the epidemiological scenario considered; see below). For scenarios with weekly SARS-CoV-2 introductions, infected patients and staff are always randomly selected in this way.

For (ii), durations of each stage of infection are drawn randomly from their respective probability distributions as soon as an Individual becomes infected.

For (iii), when a pair of Infectious and Susceptible Individuals are in contact with one another, a transmission event occurs if the calculated probability of transmission (*p* × *d)* is greater than or equal to a number randomly drawn from a uniform distribution bounded by 0% and 100%.

Although the contact network was generated stochastically as described above, the same contact network was used for each epidemic simulation, and hence variation between epidemic simulations does not result from stochasticity in inter-individual contacts.

#### Details

##### Initialization

This section describes initialization conditions for running of the model, as well as the various scenarios and sensitivity analyses considered to account for model uncertainty.

Before the first time-step in each simulation, one randomly selected Individual is infected with non-symptomatic SARS-CoV-2 infection (with equal probabilities of their infection stage being exposed, infectious pre-symptomatic, or infectious asymptomatic). This first infection is referred to as the ‘index case’. Index cases are conceived as being either infected patients newly admitted to the LTCF upon transfer from another setting, or staff who acquired infection in the community.

The following describes how initialization conditions are adjusted in the model to allow simulation of various scenarios: (i) five distinct scenarios of SARS-CoV-2 introduction into the LTCF, (ii) the baseline 170-bed LTCF vs. a smaller 30-bed LTCF geared towards elder care, and (iii) three distinct transmission rates.

##### Epidemiological scenarios of SARS-CoV-2 introduction

Five distinct epidemiological scenarios are considered, each describing a different source and frequency of SARS-CoV-2 introduction(s) into the LTCF: (baseline scenario 1, weekly patient or staff) either one infected patient admitted or one staff member infected once weekly, assuming 50% probability of patient or staff each week; (scenario 2, single patient transfer) one infected patient admitted at *t*_0_; (scenario 3, weekly patient transfer), a different infected patient admitted once weekly; (scenario 4, single infected staff), one staff member infected in the community at *t*_0_; and lastly (scenario 5, weekly infected staff) a different staff member infected in the community once weekly.

##### Two distinct LTCFs

Two distinct LTCFs are considered. The baseline LTCF uses all Individuals in the LTCF from the i-Bird study as described above. The second LTCF excludes all Individuals from Wards 1, 2, 3 and 4, leaving only individuals in Ward 5 and the Other ward. For this second LTCF, a distinct contact network was simulated in the same way as described above for the baseline LTCF. All of the five epidemiological scenarios above were also run for this second LTCF.

##### SARS-CoV-2 transmission rate

To account for uncertainty in SARS-CoV-2 transmissibility, the model for the baseline LTCF and introduction scenario is also run using extreme *R*_0_ estimates (*R*_0_=1.5, *R*_0_=6) to derive the probability of transmission per minute spent in contact (*p*=0.07%, *p*=0.21%).

#### Input

Three input files are needed to run the IBM (admission, contact and parameter files). The admission file lists dates of hospital arrival and departure for all individuals included in a simulation. The contact file lists all contacts that occur between individuals (patients and staff) over time during simulations. Here, simulated contact files were obtained as described above. The parameter file gathers all parameter values that the IBM needs.

A separate file containing parameter values was used as input to define all parameter values for Individuals. Parameter values were derived from the original contact network from the i-Bird study, including patient demographic information. However, parameters related to infection were not included, because there were simulated and modified over the course of simulations. (described below).

### III. Description of the COVID-19 surveillance algorithm

#### Overview

A surveillance algorithm was developed to distribute nasopharyngeal swabs and RT-PCR tests to Individuals from the individual-based transmission model described above, and to test them for active COVID-19 infection. Swabs and tests were distributed once per day using clinical and demographic indicators and assuming a daily limit (capacity) to the number of swabs and tests available. The 14 surveillance strategies considered differ according to which individuals were selected each day for testing. These strategies are outlined further in the main text, although more information for group testing specifically is provided below.

#### Surveillance indicators

Included indicators for surveillance were: LTCF admission (for patients only), type of individual (patient, HCW or ancillary staff), any COVID-like symptoms and severe COVID-like symptoms. Output files from epidemic simulations contained a table of all Individuals in the model, their unique ID number and their identity (patient or type of staff). For each day of each simulation, LTCF admission status and infection status for each Individual were also included. Surveillance indicators were obtained from these output files, and were supplemented with additional symptom data as described below.

#### Accounting for non-COVID but COVID-like symptoms

We generated 100 ‘symptom incidence files’ for each epidemic simulation. Symptom incidence files indicate the first day that COVID-like symptoms emerged for each Individual, and their severity (mild or severe). Note that many Individuals never experienced COVID-19 symptoms.

Since actual COVID-19 symptoms were taken from output files from epidemic simulations, they were identical across all 100 symptom incidence files within each epidemic simulation. The daily incidence of non-COVID but COVID-like symptoms (approximately 1.1% of Individuals, see Table A2) was then used to assign additional COVID-like symptoms (mild or severe) to randomly selected Individuals on each day. These non-COVID but COVID-like symptoms thus varied across each of the 100 symptom incidence files.

#### Running the surveillance algorithm

Starting on the first day of each epidemic simulation (t=0), the surveillance algorithm was run to (i) identify which individuals were to be tested, (ii) obtain test results, and (iii) to count the cumulative number of swabs and tests used. Individuals were selected randomly among those indicated if on a given day there were more Individuals indicated for testing than there were tests available. The algorithm continued until the first positive test result was returned, to a maximum of 21 days, after which all outbreaks were assumed to be detected.

#### Defining group testing

For group testing, clinical specimens (swabs) from multiple individuals were pooled and tested as one (up to, but not necessarily reaching a maximum of 2, 4, 8, 16, 32 or 64 specimens per test). SARS-CoV-2 group testing comes at the cost of reduced test sensitivity: using standard RT-PCR, Yelin *et al*. estimated a false negative rate of 10% (sensitivity=0.9) when pooling a single positive SARS-CoV-2-positive sample with 31 negative samples, all from nasal/throat swabs.(20) Conservatively assuming a 0% false negative rate in their un-pooled samples (sensitivity=1), and for simplicity assuming a linear decrease in sensitivity with each additional true negative sample, we calculated a sensitivity discounting rate per additional true negative sample:

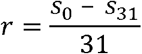

where *s*_0_ is the test sensitivity when no true negative samples were pooled with the original true positive sample, *s_31_* is the sensitivity when 31 true negative samples were pooled with the positive sample, and solving yields r=3.23×10^−3^. Hence for a group test with S specimens and P true positive specimens, we calculated sensitivity as:

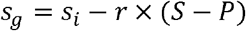

where *s_g_* and *s_i_*, are group- and individual-level test sensitivities, respectively.

Test sensitivity varied for individuals in the Exposed and Infectious stages of infection. When pooled samples included true positive swabs from both Exposed and Infectious Individuals, test sensitivity was calculated using only the Infectious Individual(s), while Exposed swabs were counted as negative samples. As with individual RT-PCR testing for SARS-CoV-2, group testing is highly specific,(24) so perfect specificity was assumed.

## **Appendix B:** Supplementary results for the article *Optimizing COVID-19 surveillance in long-term care: a modelling study*

**Table B1.**
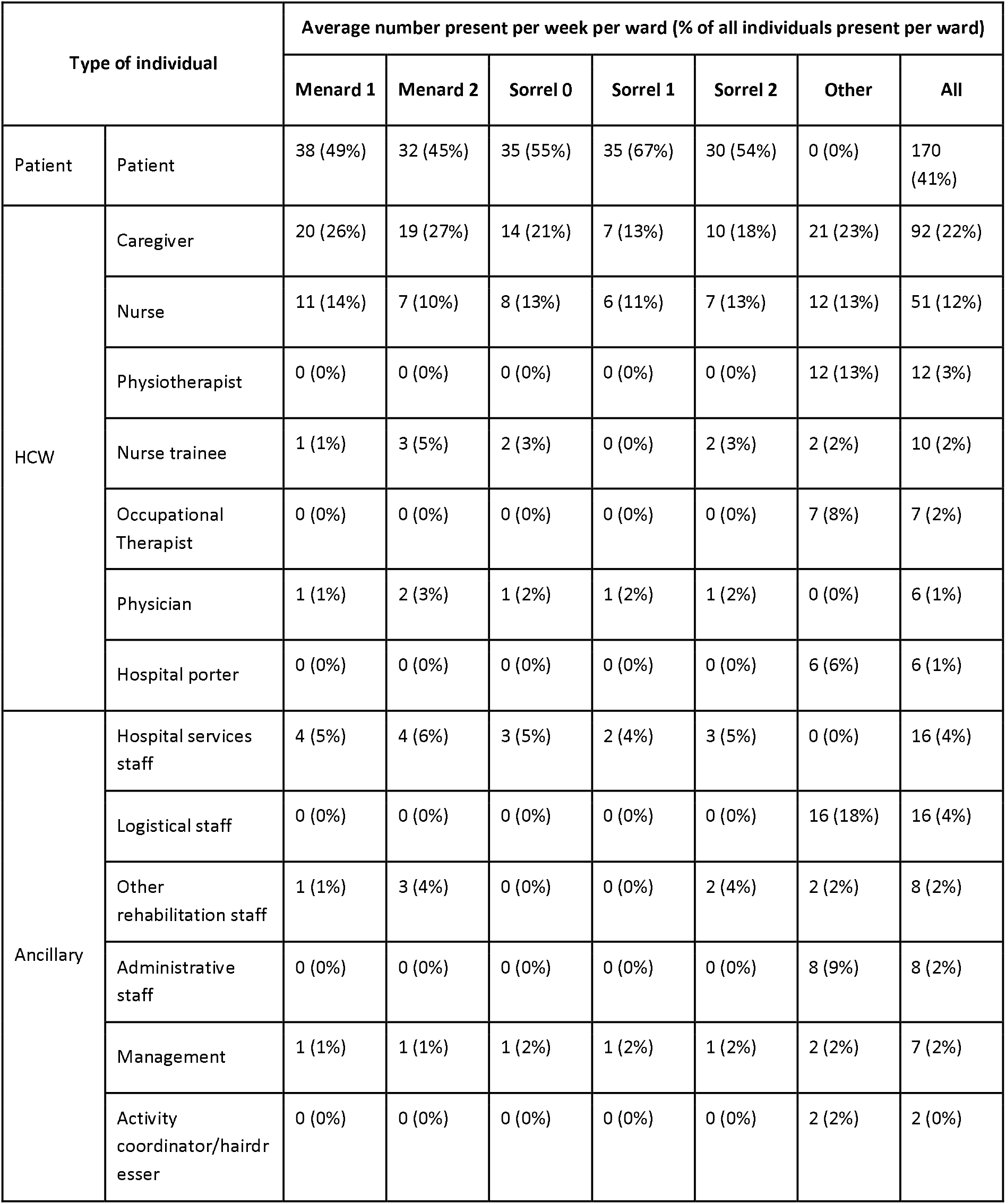
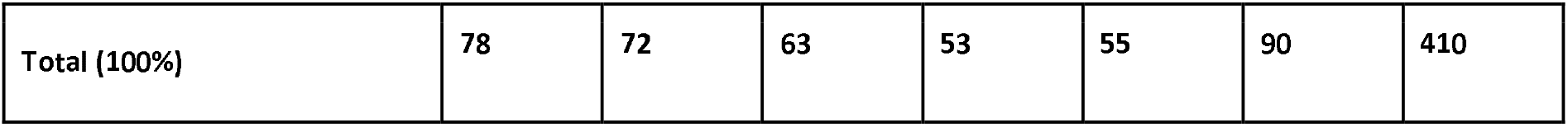
Demographic breakdown of patients and staff present in each ward of the simulated baseline LTCF. Staff were grouped as healthcare workers (HCWs) or ancillary staff. The ward ‘Other’ accounts for staff not affiliated with any one particular ward, including those who work in the back office or regularly move between wards. The geriatric ward (Sorrel 2, with its distinct patients, staff and within-ward contact network) was used independently in a sensitivity analysis to simulate outbreaks in a separate 30-bed geriatric LTCF.

**Table B2.** Simulated cumulative COVID-19 case counts over time, stratified by epidemiological scenario, LTCF, and type of individual. Only simulations resulting in outbreaks were included (for LTCF 1, 64% of simulations from scenario 4, 100% from other scenarios; for LTCF 2, 96% from scenario 2, 24% from scenario 4, 100% from other scenarios).

| Scenario | Type of individual | Median cumulative number infected per outbreak by time $t$ (95% UI) | | | | | |
| --- | --- | --- | --- | --- | --- | --- | --- |
|  |  | LTCF 1: Rehabilitation hospital (170 beds) |  |  | LTCF 2: Geriatric LTCF (30 beds) |  |  |
|  |  | 1 week (t=7) | 2 weeks (t=14) | 3 weeks (t=21) | 1 week (t=7) | 2 weeks (t=14) | 3 weeks (t=21) |
| Scenario 1: Weekly patient or staff | Patient | 3 (0 - 16) | 21 (0 - 84) | 64 (2 - 142) | 1 (0 - 10) | 7 (0 - 27) | 24 (0 - 31) |
|  | HCW | 1 (0 - 5) | 5 (0 - 25) | 22 (1 - 77) | 1 (0 - 2) | 2 (0 - 12) | 8 (1 - 18) |
|  | Ancillary staff | 0 (0 - 1) | 1 (0 - 5) | 3 (0 - 13) | 0 (0 - 1) | 1 (0 - 3) | 2 (0 - 5) |
|  | **Total** | **4 (1 - 20)** | **30 (2 - 100)** | **86 (6 - 224)** | **2 (1 - 12)** | **10 (2 - 39)** | **34 (3 - 52)** |
| Scenario 2: Single patient transfer | Patient | 7 (2 - 19) | 30 (9 - 86) | 96 (33 - 145) | 3 (1 - 11) | 19 (1 - 26) | 27 (3 - 31) |
|  | HCW | 1 (0 - 5) | 8 (1 - 25) | 32 (7 - 68) | 0 (0 - 3) | 5 (0 - 11) | 12 (1 - 17) |
|  | Ancillary staff | 0 (0 - 2) | 1 (0 - 4) | 4 (0 - 11) | 0 (0 - 1) | 0 (0 - 2) | 2 (0 - 5) |
|  | **Total** | **9 (2 - 23)** | **40 (10 - 111)** | **133 (39 - 221)** | **4 (1 - 13)** | **25 (2 - 36)** | **40 (7 - 49)** |
| Scenario 3: Weekly patient transfer | Patient | 7 (2 - 19) | 37 (11 - 93) | 109 (43 - 148) | 4 (1 - 11) | 17 (2 - 26) | 28 (12 - 32) |
|  | HCW | 1 (0 - 4) | 9 (2 - 28) | 36 (11 - 76) | 0 (0 - 4) | 4 (0 - 12) | 11 (1 - 18) |
|  | Ancillary staff | 0 (0 - 2) | 1 (0 - 4) | 5 (0 - 12) | 0 (0 - 1) | 0 (0 - 3) | 2 (0 - 5) |
|  | **Total** | **9 (2 - 21)** | **48 (15 - 116)** | **150 (62 - 227)** | **4 (1 - 13)** | **23 (3 - 38)** | **42 (13 - 51)** |
| Scenario 4: Single infected staff | Patient | 0 (0 - 6) | 3 (0 - 41) | 25 (0 - 97) | 0 (0 - 4) | 4 (0 - 22) | 18 (0 - 29) |
|  | HCW | 1 (0 - 4) | 3 (1 - 14) | 8 (1 - 43) | 1 (0 - 2) | 2 (1 - 7) | 6 (2 - 13) |
|  | Ancillary staff | 0 (0 - 1) | 0 (0 - 3) | 1 (0 - 6) | 0 (0 - 1) | 0 (0 - 1) | 1 (0 - 3) |

|  | **Total** | **2 (1 - 9)** | **6 (2 - 57)** | **36 (2 - 146)** | **2 (1 - 6)** | **6 (2 - 29)** | **26 (2 - 44)** |
| --- | --- | --- | --- | --- | --- | --- | --- |
| Scenario 5:<br>Weekly infected staff | Patient | 0 (0 - 11) | 2 (0 - 48) | 20 (0 - 108) | 0 (0 - 4) | 0 (0 - 19) | 0 (0 - 29) |
|  | HCW | 1 (0 - 4) | 3 (1 - 15) | 10 (1 - 40) | 1 (0 - 2) | 2 (0 - 7) | 3 (1 - 13) |
|  | Ancillary staff | 0 (0 - 1) | 1 (0 - 3) | 1 (0 - 6) | 0 (0 - 1) | 1 (0 - 2) | 1 (0 - 4) |
|  | **Total** | **2 (1 - 13)** | **6 (2 - 64)** | **34 (3 - 150)** | **1 (1 - 6)** | **2 (2 - 25)** | **4 (3 - 44)** |

**Table B3.**
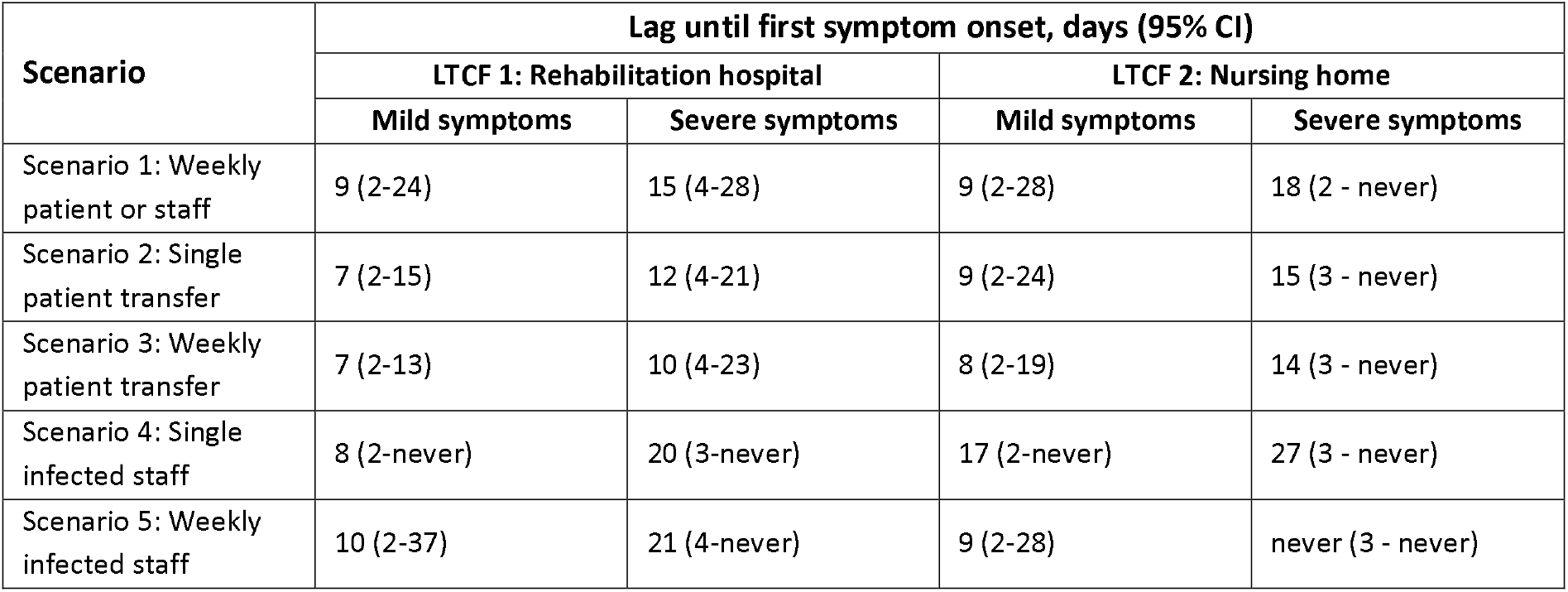
Lags between introduction of the index case and first presentation of mild and severe COVID-19 symptoms. Results are stratified by epidemiological scenario and LTCF. Only simulations resulting in outbreaks were included (for LTCF 1, 64% of simulations from scenario 4, 100% from other scenarios; for LTCF 2, 96% from scenario 2, 24% from scenario 4, 100% from other scenarios). Lags for outbreaks in which symptoms never appeared are indicated as ‘never’.

| Scenario | Lag until first symptom onset, days (95% CI) |  |  |  |
| --- | --- | --- | --- | --- |
|  | LTCF 1: Rehabilitation hospital |  | LTCF 2: Nursing home |  |
|  | Mild symptoms | Severe symptoms | Mild symptoms | Severe symptoms |
| Scenario 1: Weekly patient or staff | 9 (2-24) | 15 (4-28) | 9 (2-28) | 18 (2 - never) |
| Scenario 2: Single patient transfer | 7 (2-15) | 12 (4-21) | 9 (2-24) | 15 (3 - never) |
| Scenario 3: Weekly patient transfer | 7 (2-13) | 10 (4-23) | 8 (2-19) | 14 (3 - never) |
| Scenario 4: Single infected staff | 8 (2-never) | 20 (3-never) | 17 (2-never) | 27 (3 - never) |
| Scenario 5: Weekly infected staff | 10 (2-37) | 21 (4-never) | 9 (2-28) | never (3 - never) |

**Table B4.**
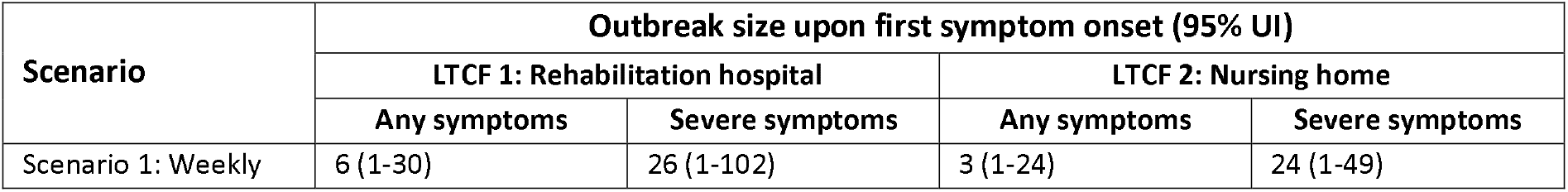
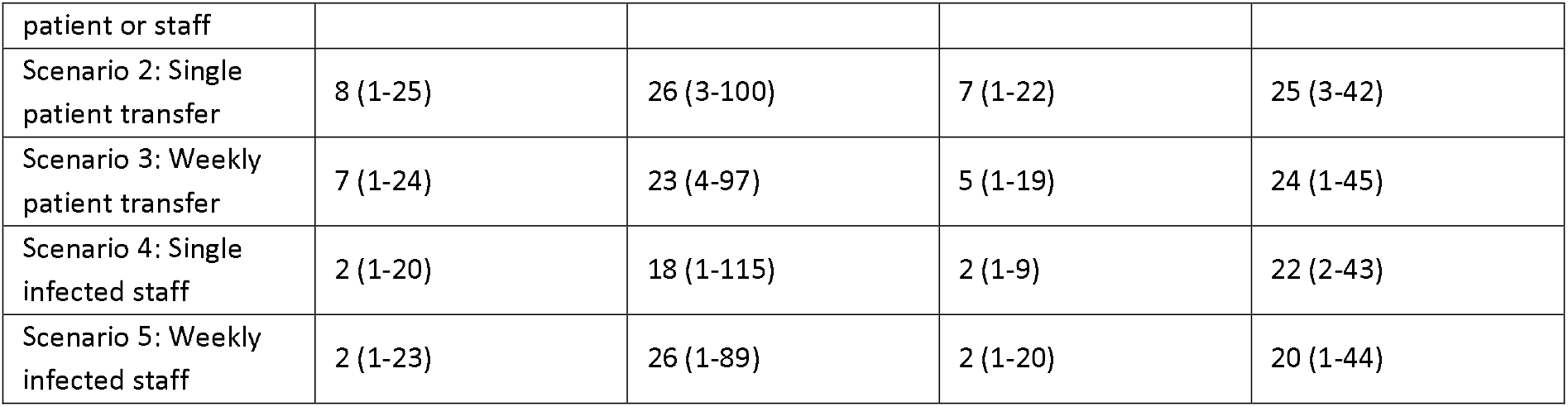
The cumulative number of individuals infected on the day that COVID-19 symptoms first appeared among anyone present in the LTCF (outbreak size upon first symptom onset). Results are stratified by epidemiological scenario and LTCF. Only simulations resulting in outbreaks were included (for LTCF 1, 64% of simulations from scenario 4, 100% from other scenarios; for LTCF 2, 96% from scenario 2, 24% from scenario 4, 100% from other scenarios), and outbreaks were excluded if no symptoms ever occurred over the course of the outbreak.

**Figure B1.**
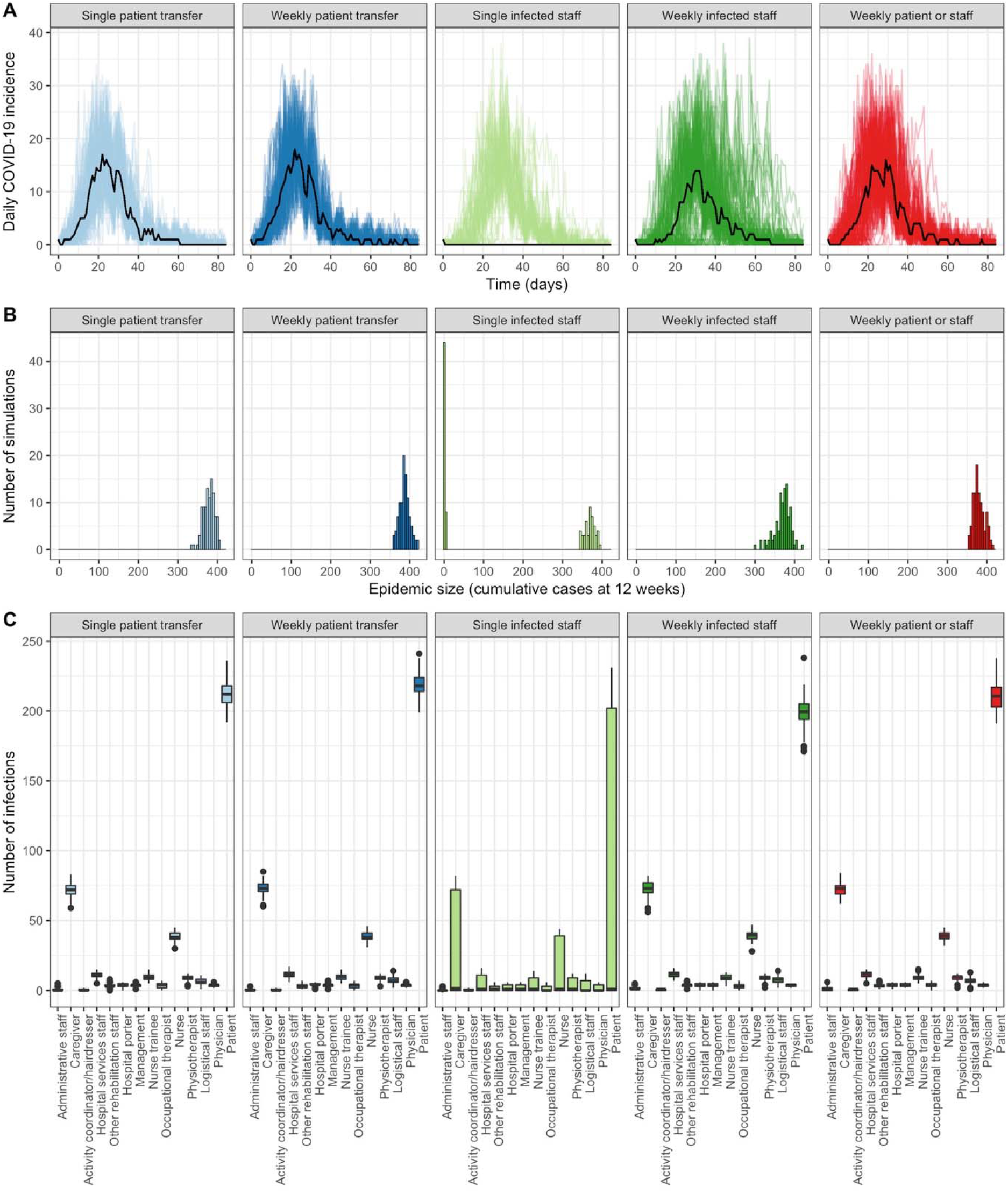
Epidemiological characteristics of COVID-19 epidemics simulated over a 12-week period in the baseline LTCF, in the absence of any surveillance, control measures or interventions, and comparing different scenarios of SARS-CoV-2 into the LTCF (columns). **A)** The daily incidence of COVID-19 infection among all patients and staff, with each coloured line representing a different simulation. Black lines represent the median daily incidence across all simulations. **B)** Histograms of the final epidemic size at 12 weeks (NB: data are naturally censured by the 12-week simulation period). **C)** Distributions of cumulative infection totals at 12 weeks among the fourteen different categories of individuals present in the hospital.

**Figure B2.**
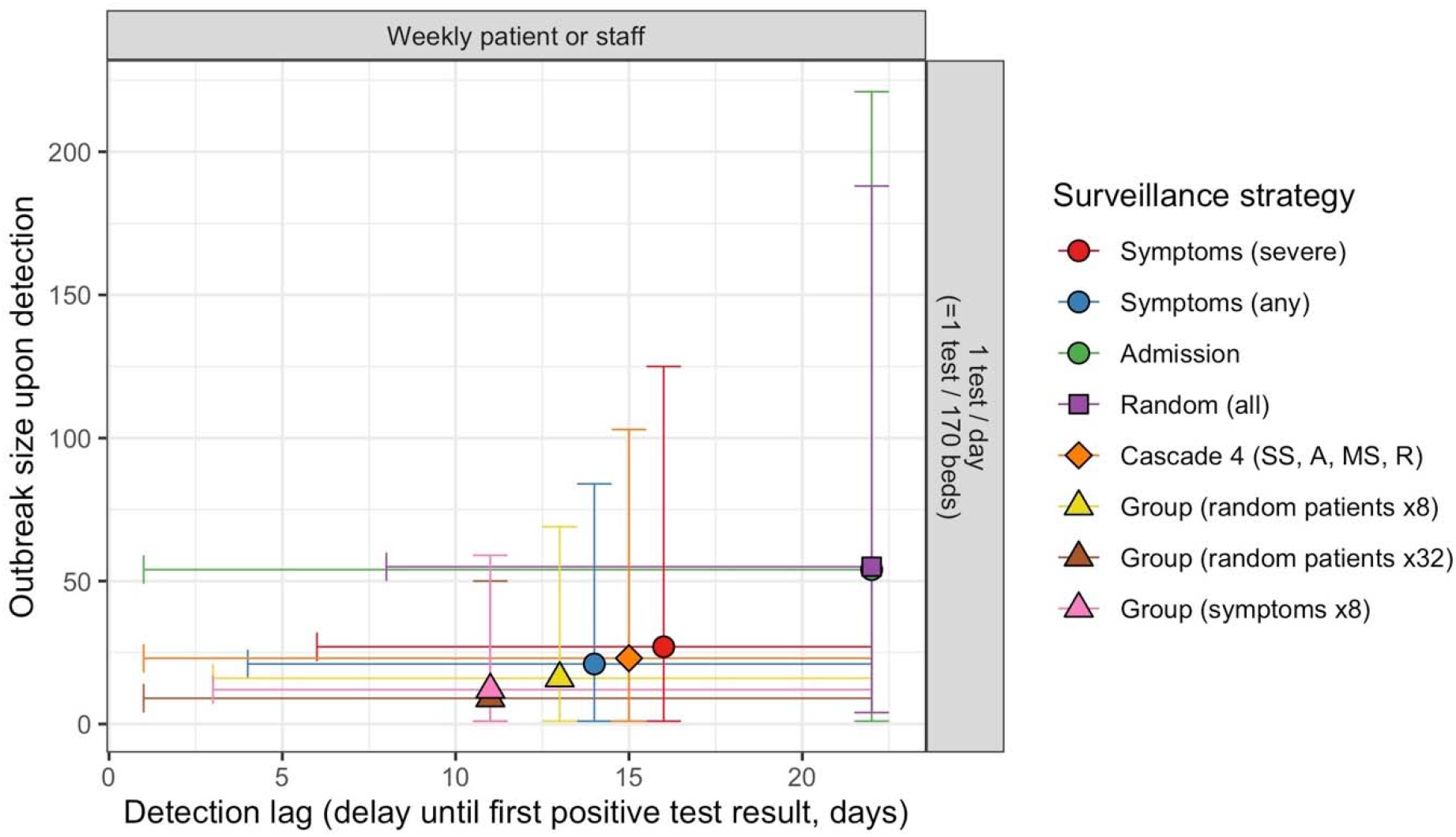
Relationship between detection lag (x-axis) and the number of undetected cases upon outbreak detection (y-axis), for selected surveillance strategies (colours), for the baseline SARS-CoV-2 importation scenario and at a testing capacity of 1 test/day. Symbols represent medians and error bars represent 95% credible intervals.

**Figure B3.**
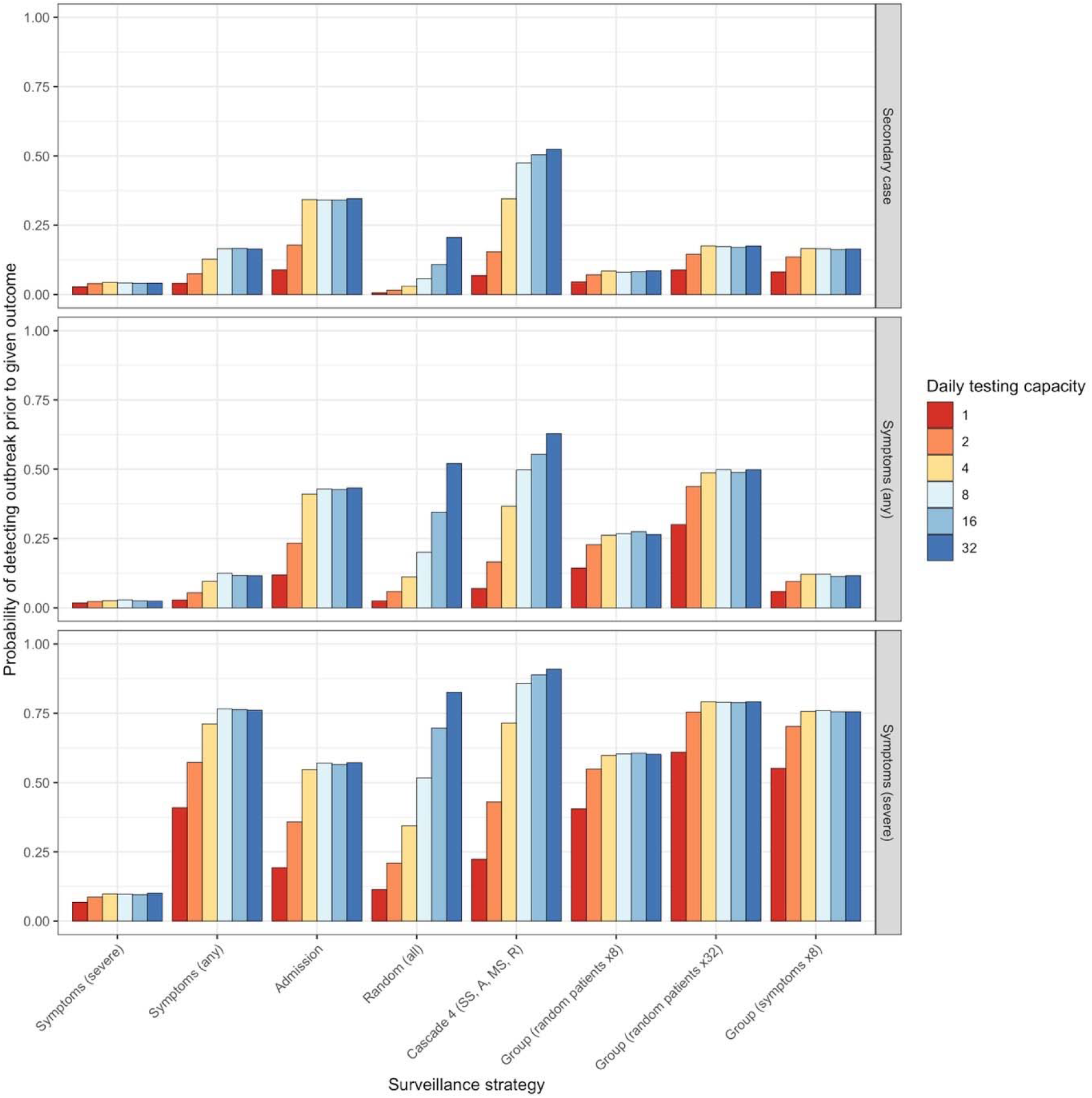
Probability of detecting COVID-19 outbreaks before any secondary cases (top panel), before the onset of any COVID-19 symptoms (middle panel), and before the onset of any severe COVID-19 symptoms (bottom panel). Probabilities depended on the surveillance strategy considered (x-axis) and the daily testing capacity (colours), and saturate at high testing capacity for all but random and cascade strategies.

**Figure B4.**
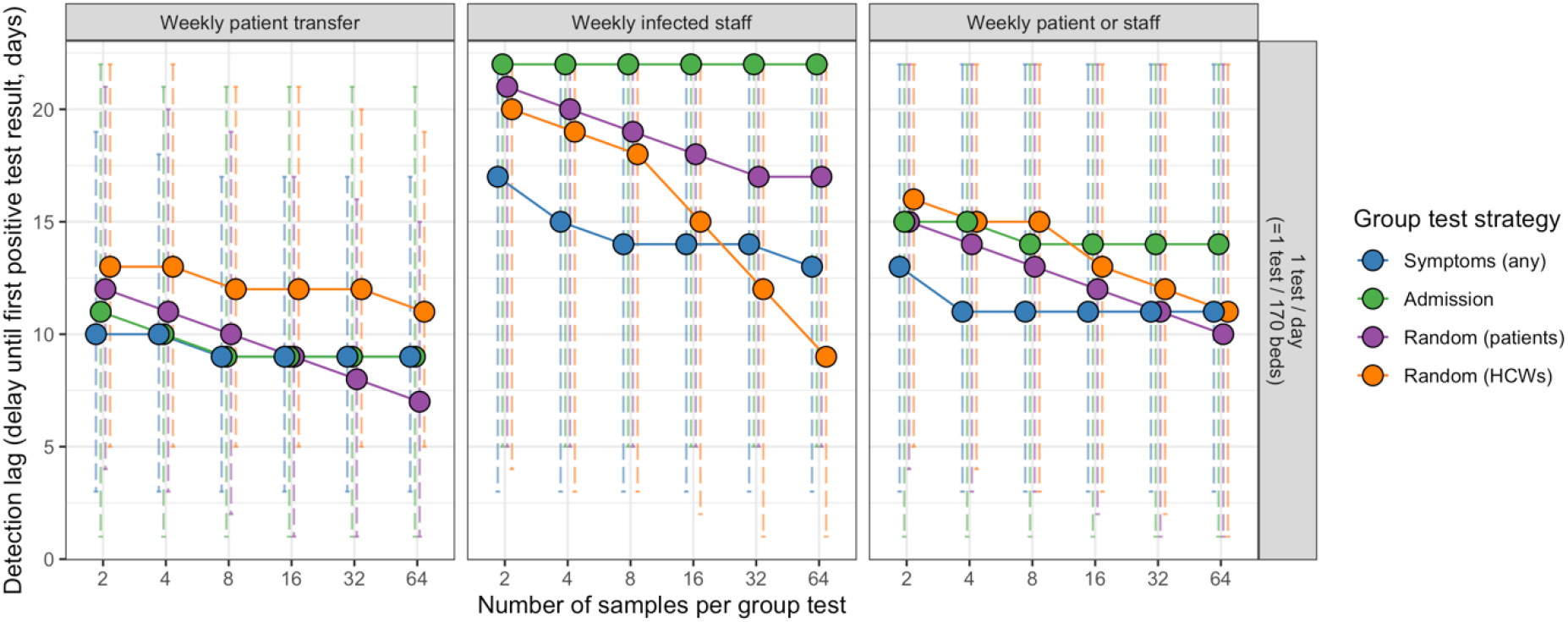
Efficacy of group testing strategies depend on the which individuals are indicated for inclusion in group tests (colours), the maximum number of specimens potentially included per group test (x-axis), and the epidemiological scenario considered (columns). In the main analysis, the number of samples per group test was capped at 32, owing to uncertainty about sufficient test sensitivity above this threshold. Circles represent medians and error bars represent 95% uncertainty intervals.

**Figure B5.**
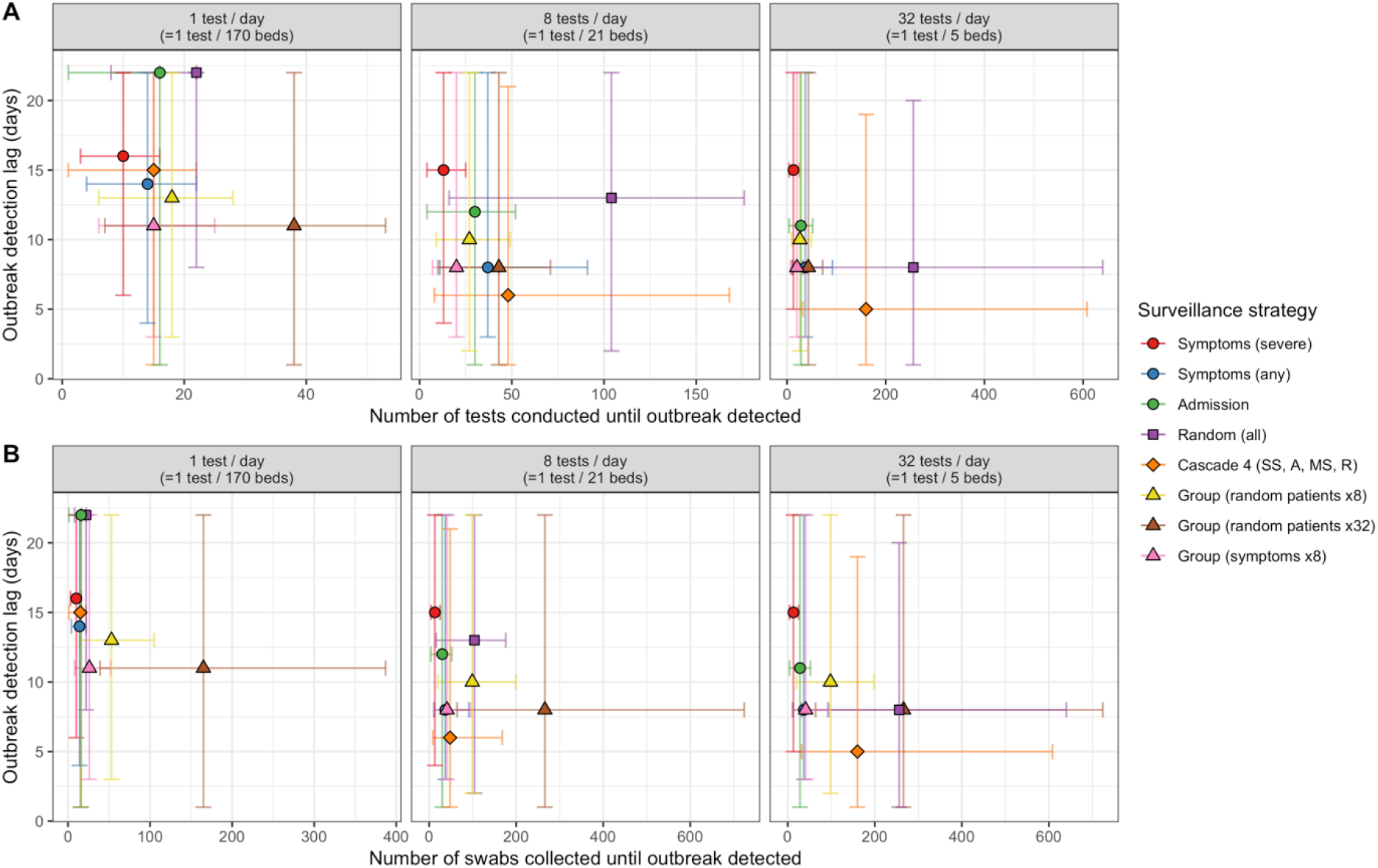
Efficiency plots for selected surveillance strategies, comparing the efficacy (y-axis) and resource use (x-axis) in terms of the number of tests used (top row) and swabs collected (bottom row) until outbreaks were detected. The assumed daily testing capacity varies across columns. Symbols represent medians and error bars represent 95% uncertainty intervals across all outbreak simulations. Overlapping symbols were shifted along the x-axis by up to 5 units and can be identified by reduced size of error bar whiskers. For cascades: SS=severe symptoms, MS=mild symptoms, A=admission, R=random (patients).

**Figure B6.**
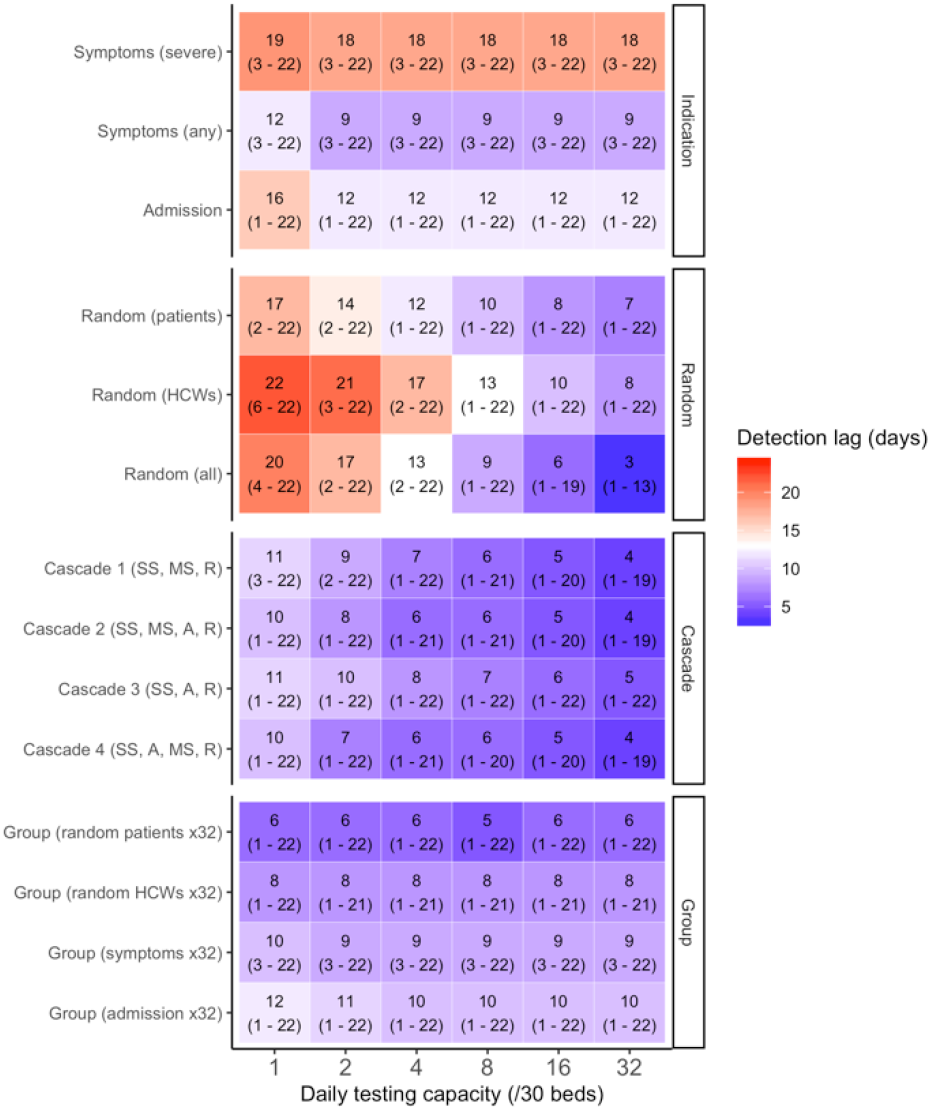
For simulations in the 30-bed geriatric LTCF, median lags to outbreak detection (95% uncertainty interval) are shown for each surveillance strategy (y-axis) as a function of the daily testing capacity (x-axis). Group testing strategies assume a maximum of 32 swabs per test.

**Figure B7.**
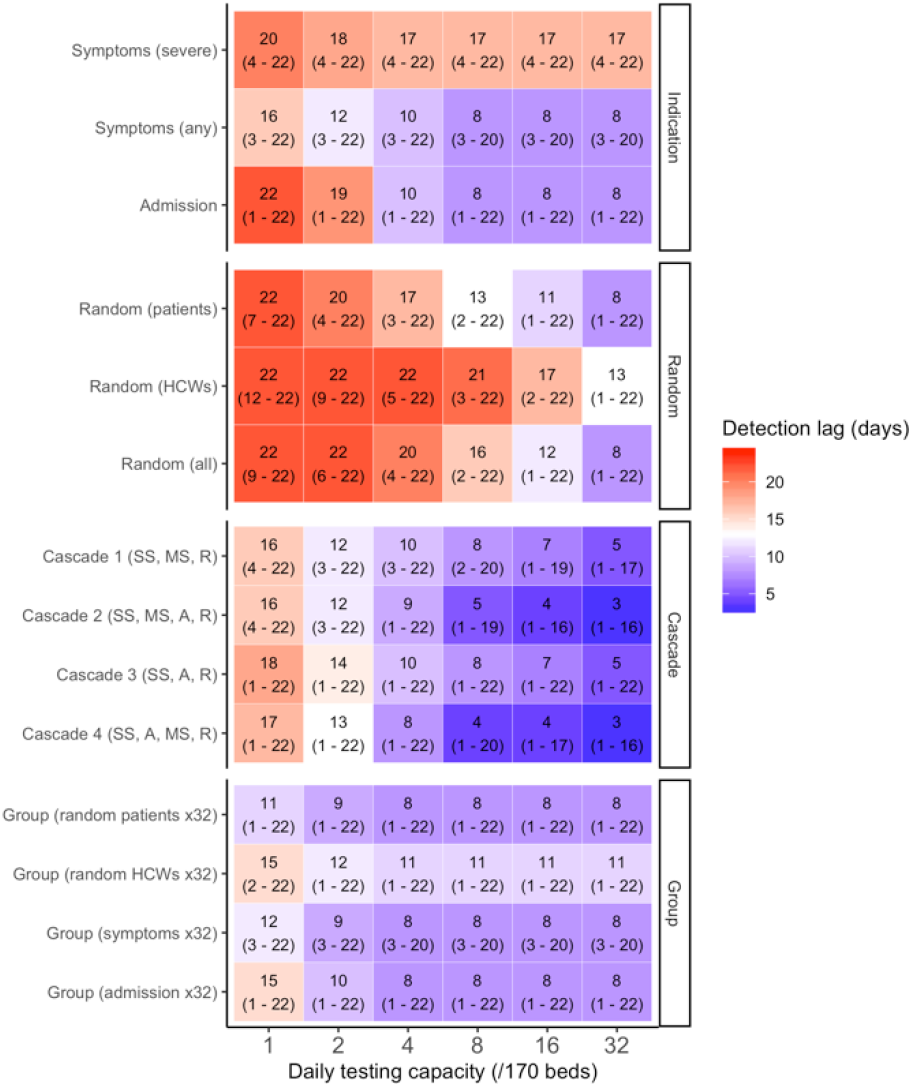
For simulations in the baseline LTCF with a low transmission rate (*p*=0.07%), median lags to outbreak detection (95% uncertainty interval) are shown for each surveillance strategy (y-axis) as a function of the daily testing capacity (x-axis). Group testing strategies assume a maximum of 32 swabs per test.

**Figure B8.**
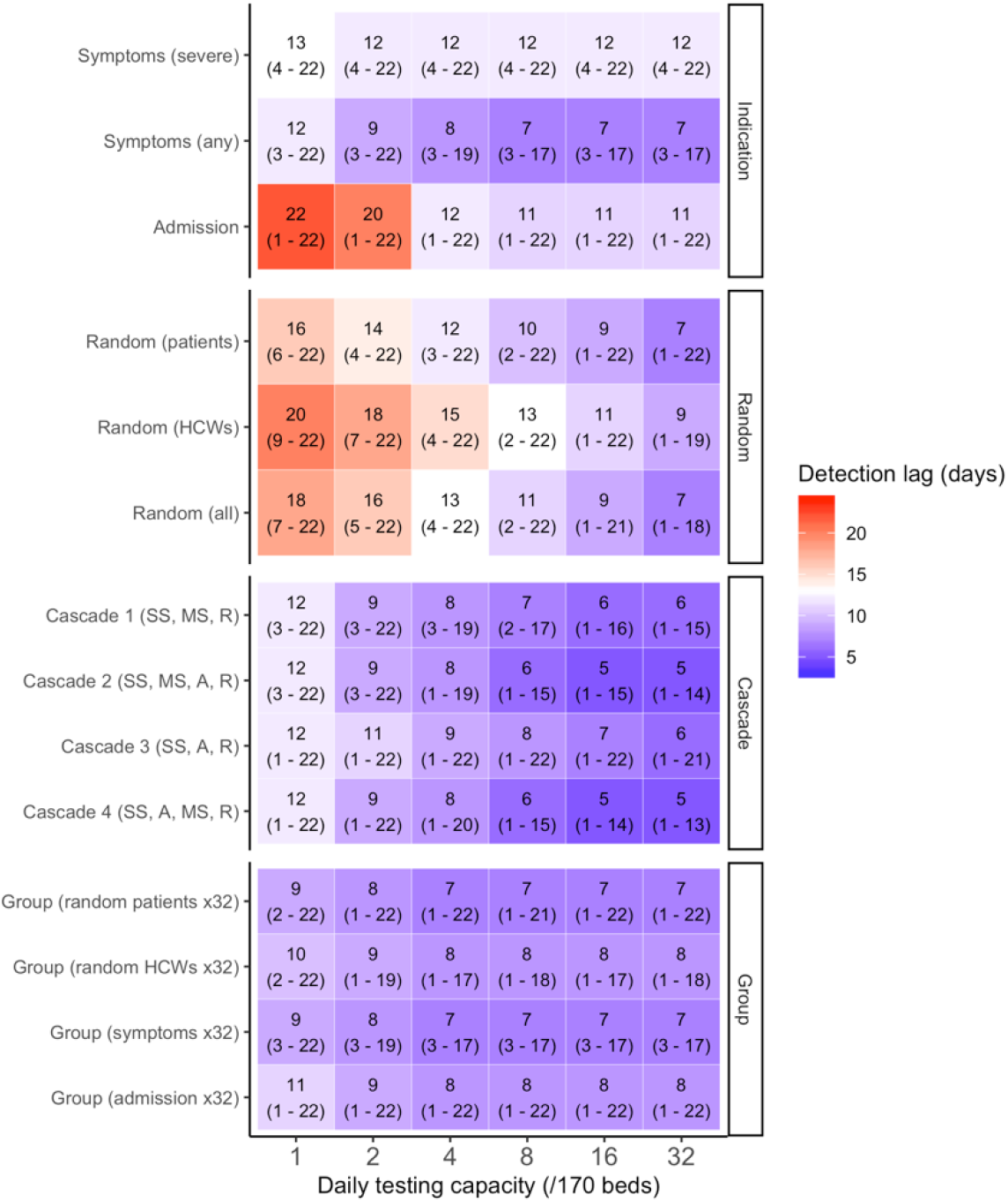
For simulations in the baseline LTCF with a high transmission rate (p=0.21%), median lags to outbreak detection (95% uncertainty interval) are shown for each surveillance strategy (y-axis) as a function of the daily testing capacity (x-axis). Group testing strategies assume a maximum of 32 swabs per test.

**Figure B9.**
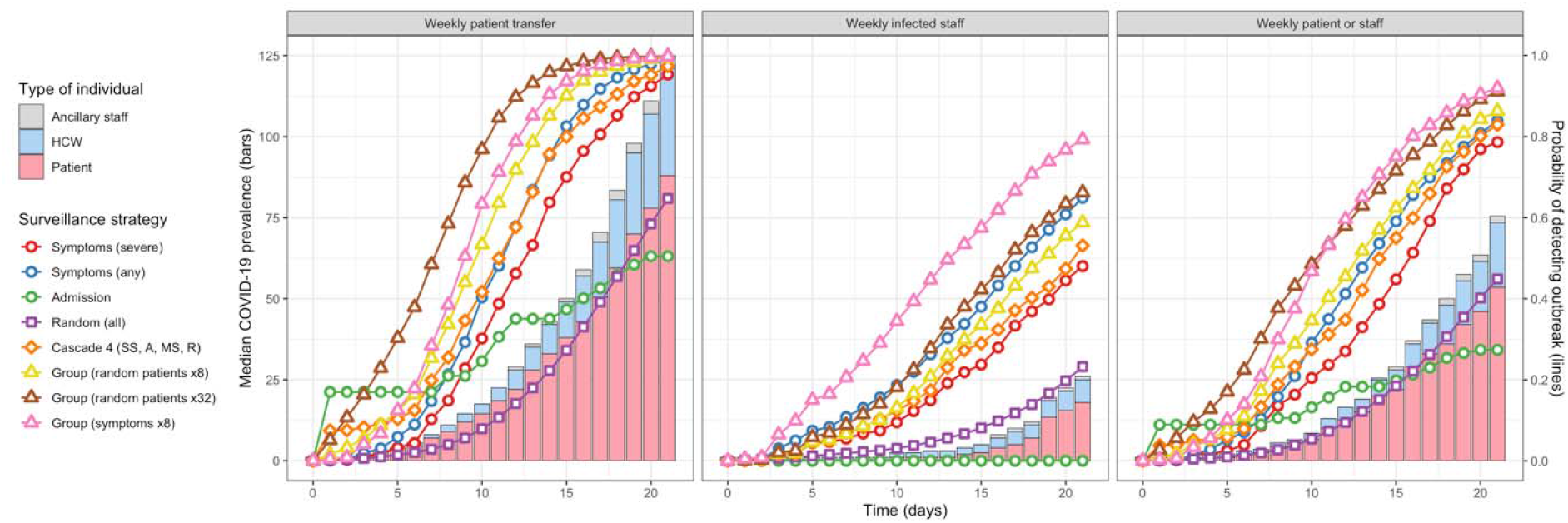
The size of COVID-19 outbreaks varied over time (vertical bars) across SARS-CoV-2 importation scenarios (columns). Consequently, the probability of detecting outbreaks over time using different surveillance strategies (coloured lines) varied as a result of differences in how many, and which types of individuals became infected over time across these different importation scenarios; here, testing capacity = 1 test/day.

**Figure B10.**
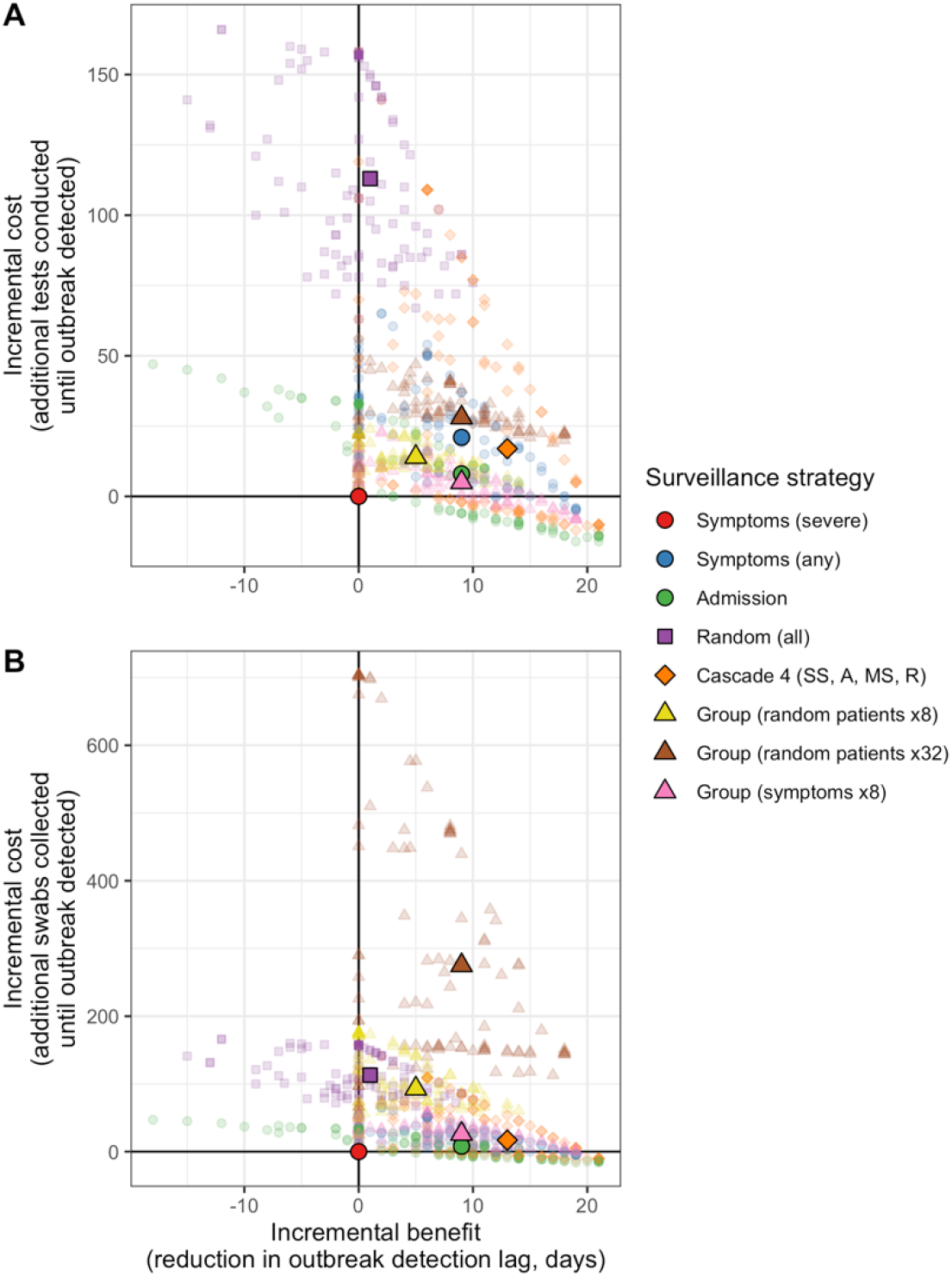
Incremental efficiency plots for simulations in the baseline LTCF with a low transmission rate (*p*=0.07%). Daily testing capacity is fixed at 8 tests/day, small translucent points represent median outcomes across all surveillance simulations for each simulated outbreak, and larger opaque points represent median outcomes across all outbreaks.

**Figure B11.**
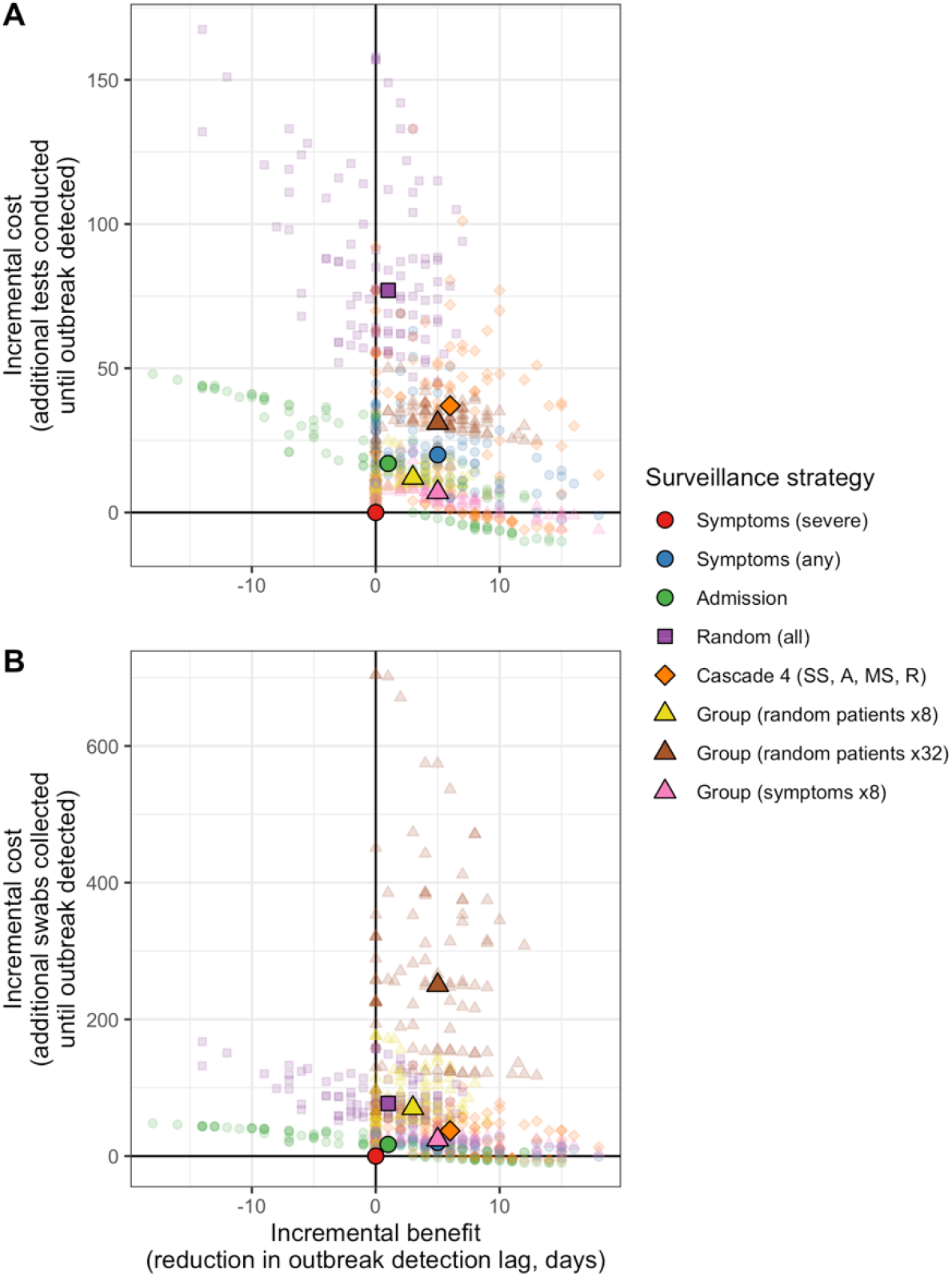
Incremental efficiency plots for simulations in the baseline LTCF with a high transmission rate (*p*=0.21%). Daily testing capacity is fixed at 8 tests/day, small translucent points represent median outcomes across all surveillance simulations for each simulated outbreak, and larger opaque points represent median outcomes across all outbreaks.

**Figure B12.**
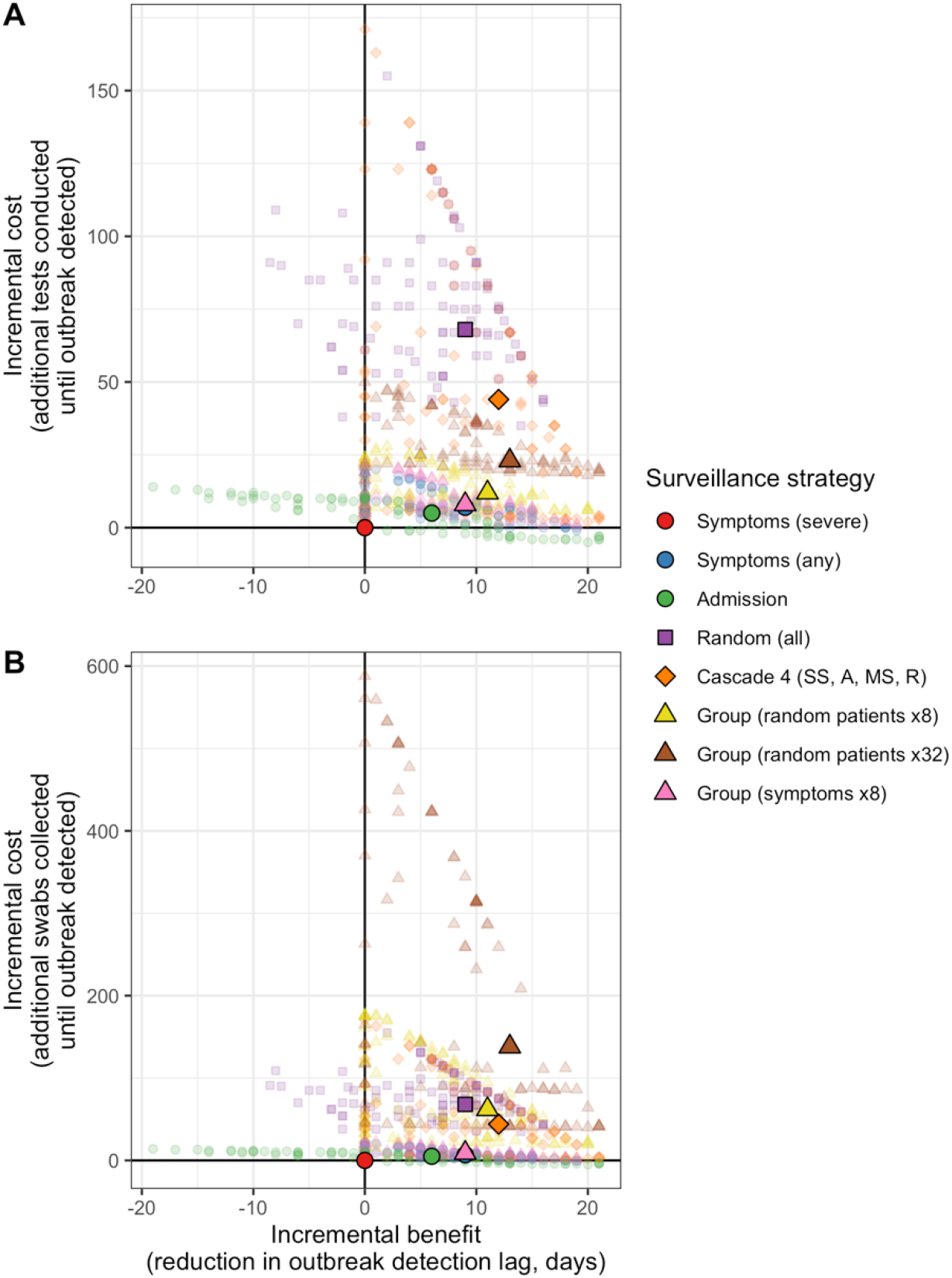
Incremental efficiency plots for simulations in the 30-bed geriatric LTCF. Daily testing capacity is fixed at 8 tests/day, small translucent points represent median outcomes across all surveillance simulations for each simulated outbreak, and larger opaque points represent median outcomes across all outbreaks.

